# The role of adolescent lifestyle habits in biological aging: A prospective twin study

**DOI:** 10.1101/2022.05.30.22275761

**Authors:** Anna Kankaanpää, Asko Tolvanen, Aino Heikkinen, Jaakko Kaprio, Miina Ollikainen, Elina Sillanpää

## Abstract

Adolescence is a stage of fast growth and development. Exposures during puberty can have long-term effects on health in later life. This study aims to investigate the role of adolescent lifestyle in biological aging. The study participants originated from the longitudinal FinnTwin12 study (n = 5114). Adolescent lifestyle-related factors, including body mass index (BMI), leisure-time physical activity, smoking and alcohol use, were based on self-reports and measured at ages 12, 14 and 17 years. For a subsample, blood-based DNA methylation (DNAm) was used to assess biological aging with six epigenetic aging measures in young adulthood (21–25 years, n = 824). A latent class analysis was conducted to identify patterns of lifestyle behaviors in adolescence, and differences between the subgroups in later biological aging were studied. Genetic and environmental influences on biological aging shared with lifestyle behavior patterns were estimated using quantitative genetic modelling.

We identified five subgroups of participants with different adolescent lifestyle behavior patterns. When DNAm GrimAge, DunedinPoAm and DunedinPACE estimators were used, the class with the unhealthiest lifestyle and the class of participants with high BMI were biologically older than the classes with healthier lifestyle habits. The differences in lifestyle-related factors were maintained into young adulthood. Most of the variation in biological aging shared with adolescent lifestyle was explained by common genetic factors. These findings suggest that an unhealthy lifestyle during pubertal years is associated with accelerated biological aging in young adulthood. Genetic pleiotropy can largely explain the observed associations.

## INTRODUCTION

Epidemiological studies of life course have indicated that exposures during early life has long-term effects on later health (Kuh et al., 2003). Unhealthy environments and lifestyle habits during rapid cell division can affect the structure or functions of organs, tissues or body systems, and these changes can subsequently affect health and disease in later life (Power et al., 2013). For example, lower birth weight and fast growth during childhood predispose individuals to coronary heart disease and increased blood pressure in adulthood (Osmond & Barker, 2000). In addition to infancy and childhood, adolescence is also a critical period of growth.

Adolescence is characterised by pubertal maturation and growth spurts. Early pubertal development is linked to worse health conditions, such as obesity and cardiometabolic risk factors in adulthood (Prentice & Viner, 2013). However, childhood obesity can lead to early onset of puberty, especially among girls (Li et al., 2017; Richardson et al., 2020) and, therefore, can confound the observed associations between early pubertal development and worse later health. Moreover, early pubertal development is linked to substance use and other risky behaviors in adolescence (Hartman et al., 2017; Savage et al., 2018), but the associations are partly explained by familial factors (Savage et al., 2018).

Many unhealthy lifestyle choices, such as smoking initiation, alcohol use and a physically inactive lifestyle, are already made in adolescence and increase the risk of developing several non-communicable diseases over the following decades (Lopez et al., 2006). Once initiated, unhealthy habits are likely to persist into adulthood (Latvala et al., 2014; Maggs & Schulenberg, 2005; Rovio et al., 2018; Salin et al., 2019). A recent systematic review showed that healthy habits tend to cluster during childhood and adolescence (Whitaker et al., 2021). Typically, about half of the adolescents fall into subgroups characterised by healthy lifestyle habits. However, small minorities of adolescents are classified as heavy substance users or as having multiple other risk behaviors. The long-term consequences of the accumulation of unhealthy adolescent behaviors on health in later life have been rarely studied.

An unhealthy lifestyle in adolescence can affect biological mechanisms of aging at the molecular level and, subsequently, morbidity. Epigenetic alterations, including age-related changes in DNA methylation (DNAm), constitute a primary hallmark of biological aging (López-Otín et al., 2013). Epigenetic clocks are algorithms that aim to quantify biological aging using DNAm levels within specific CpG sites. The first-generation clocks, Horvath’s and Hannum’s clocks, were trained to predict chronological age (Hannum et al., 2013; Horvath, 2013), whereas the second-generation clocks, such as DNAm PhenoAge and GrimAge, are better predictors of health span and lifespan (Levine et al., 2018; Lu et al., 2019). For epigenetic clocks, the difference between an individual’s epigenetic age estimate and chronological age provides a measure of age acceleration (AA). The DunedinPoAm estimator differs from its predecessors in that it has been developed to predict the pace of aging (Belsky et al., 2020). The pace of aging describes longitudinal changes over time in several biomarkers of organ-system integrity among same-aged individuals. Recently, the DunedinPACE estimator, which constitutes an advance on the original DunedinPoAm, was published (Belsky et al., 2022). DunedinPACE measures the pace of aging more precisely and has better test-retest reliability. From the life course perspective, epigenetic aging measures are useful tools to assess biological aging at all ages and detect changes induced by lifetime exposures.

Previous studies have linked several lifestyle-related factors, such as higher body mass index (BMI), smoking, alcohol use and lower leisure-time physical activity (LTPA), with accelerated biological aging measured using epigenetic clocks (Oblak et al., 2021). However, most of these studies were based on cross-sectional data on older adults. The first studies on the associations of adolescent lifestyle-related exposures with biological aging assessed with epigenetic aging measures indicated that advanced pubertal development, higher BMI and smoking are associated with accelerated biological aging in adolescence (Etzel et al., 2021; Raffington et al., 2021; Simpkin et al., 2017).

The few previous studies conducted on this topic have focused on single lifestyle factors, and a comprehensive understanding of the role of adolescent lifestyle in later biological aging remains unclear. Our first aim is to define the types of lifestyle behavior patterns that can be identified in adolescence using data-driven latent class analysis (LCA). The second aim is to investigate whether the identified behavioral subgroups differ in biological aging in young adulthood and whether the associations are independent of baseline pubertal development.

The third aim is to assess the genetic and environmental influences shared between biological aging and adolescent lifestyle behavior patterns.

## MATERIALS AND METHODS

The participants were members of the longitudinal FinnTwin12 study (born during 1983–87) (Kaprio, 2013; Rose et al., 2019). A total of 5,600 twins and their families initially enrolled in the study. At the baseline, the twins filled out the questionnaires regarding their lifestyle-related habits at 11–12 years of age, and follow-up assessments were conducted at ages 14 and 17.5 years. The response rates were high for each assessment (85%–90%). In young adulthood, at an average age of 22 years, blood samples for DNA analyses (n = 824) were collected during in-person clinical studies after written informed consent was signed. The data on health-related behaviors were collected with questionnaires and interviews. A total of 1,295 twins of the FinnTwin12 cohort were examined and measured, either in-person or through telephonic interviews. Data collection was conducted in accordance with the Declaration of Helsinki. The Indiana University IRB and the ethics committees of the University of Helsinki and Helsinki University Central Hospital approved the study protocol.

### DNAm and assessment of biological age

DNAm profiles were obtained using Illumina’s Infinium HumanMethylation450 BeadChip or the Infinium MethylationEPIC BeadChip (Illumina, San Diego, CA, USA). A detailed description of the pre-processing and normalizing of the DNAm data is provided in the Appendix (Supplementary text).

We utilized six epigenetic clocks. The first four clocks, namely Horvath’s and Hannum’s epigenetic clocks (Hannum et al., 2013; Horvath, 2013) and DNAm PhenoAge and DNAm GrimAge estimators (Levine et al., 2018; Lu et al., 2019), produced DNAm-based epigenetic age estimates in years by using a publicly available online calculator (https://dnamage.genetics.ucla.edu/new). For these measures, AA was defined as the residual obtained from regressing the estimated epigenetic age on chronological age (AA_Horvath_, AA_Hannum_, AA_Pheno_ and AA_Grim_, respectively). The fifth and sixth clocks, namely DunedinPoAm and DunedinPACE estimators, provided an estimate for the pace of biological aging in years per calendar year (Belsky et al., 2020, 2022). DunedinPoAm and DunedinPACE were calculated using publicly available R packages (https://github.com/danbelsky/DunedinPoAm38 and https://github.com/danbelsky/DunedinPACE, respectively).

The components of DNAm GrimAge (adjusted for age) were also obtained, including DNAm-based smoking pack-years and the surrogates for plasma proteins (DNAm-based plasma proteins): DNAm adrenomedullin (ADM), DNAm beta-2-microglobulin (B2M), DNAm cystatin C, DNAm growth differentiation factor 15 (GDF15), DNAm leptin, DNAm plasminogen activator inhibitor 1 (PAI-1), and DNAm tissue inhibitor metalloproteinases 1 (TIMP-1).

### Lifestyle-related factors in adolescence

#### Body mass index (BMI) at ages 12, 14 and 17 years

BMI (kg/m^2^) was calculated based on self-reported height and weight.

#### Leisure-time physical activity (LTPA) at ages 12, 14 and 17 years

The frequency of LTPA at the age of 12 years was assessed with the question ‘How often do you engage in sports (i.e. team sports and training)?’ The answers were classified as 0 = less than once a week, 1 = once a week and 2 = every day. At ages 14 and 17 years, the question differed slightly: ‘How often do you engage in physical activity or sports during your leisure time (excluding physical education)?’ The answers were classified as 0 = less than once a week, 1 = once a week, 2 = 2–5 times a week and 3 = every day.

#### Smoking status at ages 14 and 17 years

was determined using the self-reported frequency of smoking and classified as 0 = never smoker, 1 = former smoker, 2 = occasional smoker and 3 = daily smoker.

#### Alcohol use (binge drinking) at ages 14 and 17 years

The frequency of drinking to intoxication had the following classes: ‘How often do you get really drunk?’ 0 = never, 1 = less than once a month, 2 = approximately once or twice a month and 3 = once a week or more.

#### Pubertal development at age 12 years

Baseline pubertal development was assessed using a five-item Pubertal Development Scale (PDS) questionnaire (Petersen et al., 1988). Both sexes answered three questions each concerning growth in height, body hair and skin changes.

Moreover, boys were asked questions about the development of facial hair and voice change, while girls were asked about breast development and menarche. Each question had response categories 1 = growth/change has not begun, 2 = growth/change has barely started and 3 = growth/change is definitely underway, except for menarche, which was dichotomous 1 = has not occurred or 3 = has occurred (see also Wehkalampi et al., 2008). The highest response category of the original questionnaire (development completed) was omitted for all items, except for menarche, because completing the development was assumed to be very rare by the age of 12. PDS was calculated as the mean score of the five items, and higher values indicated more advanced pubertal development at age 12 years.

#### Lifestyle-related factors in young adulthood at age 21–25 years BMI

(kg/m^2^) was calculated based on the measured height and weight.

#### LTPA

was assessed using the Baecke questionnaire (Baecke et al., 1982). A sport index was based on the mean scores of four questions on sports activity described by Baecke et al. (1982) and Mustelin et al. (2012) for the FinnTwin12 study. The sport index is a reliable and valid instrument to measure high-intensity physical activity (Richardson et al., 1995).

#### Smoking

was self-reported and classified as never, former or current smoker.

#### Alcohol use

(100% alcohol grams/day) was derived from the Semi-Structured Assessment for the Genetics of Alcoholism (Bucholz et al., 1994) and based on quantity and frequency of use and the content of alcoholic beverages, assessed by trained interviewers.

### Statistical analysis

#### Patterns of lifestyle behaviors in adolescence

To identify the patterns of lifestyle behaviors in adolescence, a latent class analysis (LCA) was conducted, which is a data-driven approach to identify homogenous subgroups in a heterogeneous population. The classification was based on BMI and LTPA at ages 12, 14 and 17 years and smoking status and alcohol use at ages 14 and 17 years (10 indicator variables). All variables were treated as ordinal variables, except for continuous BMI. The classification was based on the thresholds of the ordinal variables and the means and variances of BMI.

An LCA model with 1 to 8 classes was fitted. The following fit indices were used to evaluate the goodness of fit: Akaike’s information criterion, Bayesian information criterion and sample size-adjusted Bayesian information criterion. The lower values of the information criteria indicated a better fit for the model. Moreover, we used the Vuong–Lo–Mendell– Rubin likelihood ratio (VLMR) test and the Lo–Mendell–Rubin (LMR) test to determine the optimal number of classes. The estimated model was compared with the model with one class less, and the low *p* value suggested that the model with one class less should be rejected. At each step, the classification quality was assessed using the average posterior probabilities for most likely latent class membership (AvePP). AvePP values close to 1 indicate a clear classification. In addition to the model fit, the final model for further analyses was chosen based on the parsimony and interpretability of the classes.

#### Differences in biological aging

The mean differences in biological aging between the lifestyle behavior patterns were studied using the Bolck–Croon–Hagenaars approach (Asparouhov & Muthén, 2021). The class-specific weights for each participant were computed and saved during the latent class model estimation. After that, a secondary model conditional on the latent lifestyle behavior patterns was specified using weights as training data: Epigenetic aging measures were treated as distal outcome one at a time, and the mean differences across classes were studied while adjusting for sex, age and baseline pubertal development. Similarly, the mean differences in the components of DNAm GrimAge and lifestyle-related factors in young adulthood were studied. The models of epigenetic aging measures were additionally adjusted for BMI in adulthood. To evaluate the effect sizes, standardised mean differences (SMDs) were calculated.

#### Genetic and environmental influences

Genetic and environmental influences on biological aging in common with lifestyle behavior patterns were studied using quantitative genetic modelling. For simplicity, we adjusted the epigenetic aging variables for sex, age and baseline pubertal development prior to the analysis.

We first carried out univariate modelling to study genetic and environmental influences on epigenetic aging measures (Supplementary text) (Neale & Cardon, 1992). On one hand, total variance in biological aging was decomposed in the components explained by genetic, unshared and shared environmental factors 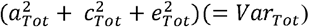 (Figure 1A). On the other hand, it comprised the variance explained by the adolescent lifestyle behavior patterns (*Var*_*Model*_) and the variance of the residual term (*Var*_*Res*_). We also conducted univariate modelling for the residual term of biological aging, which corresponds to the variation in biological aging not explained by the adolescent lifestyle behavior patterns 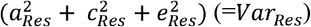 (Figure 1B). The residual terms were obtained by specifying a latent variable corresponding to the residuals of the secondary model described above (without including covariates), and the factor scores were saved. Finally, the proportion of variation in biological aging explained by the genetic factors shared with adolescent lifestyle patterns was evaluated as follows: 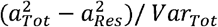. The proportion of variation in epigenetic aging explained by the environmental factors was evaluated similarly.

**Figure 1.**
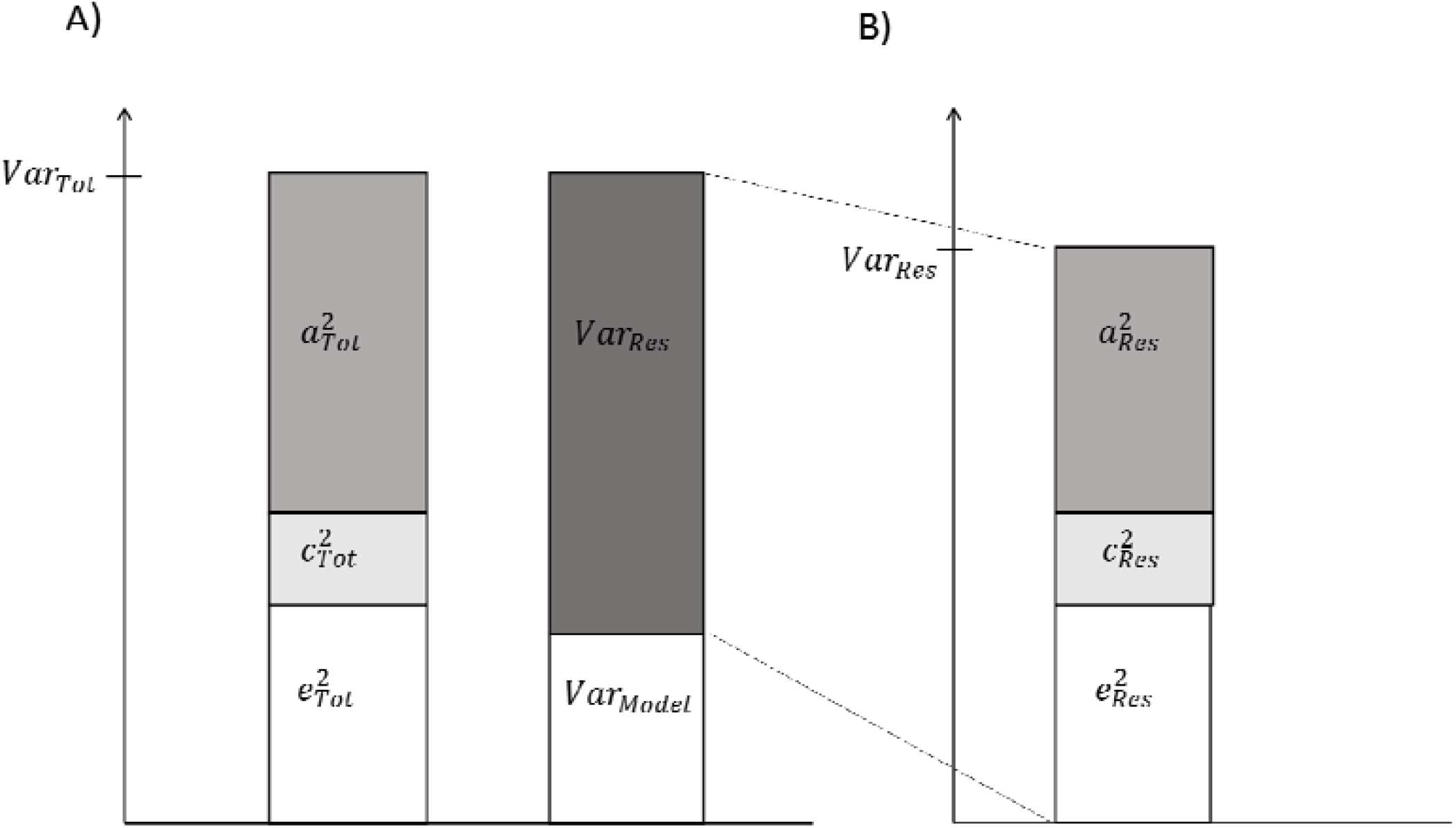
Decomposition of A) total variation in biological aging, and B) the variation of the residual term.

Missing data were assumed to be missing at random (MAR). The model parameters were estimated using the full information maximum likelihood (FIML) method with robust standard errors. Under the MAR assumption, the FIML method produced unbiased parameter estimates. The standard errors of the latent class models and secondary models were corrected for nested sampling (TYPE = COMPLEX). Descriptive statistics were calculated using IBM SPSS Statistics for Windows, version 20.0 (IBM Corp, Armonk, NY), and further modeling was conducted using Mplus, version 8.2 (Muthén & Muthén, 1998-2018).

## RESULTS

The descriptive statistics of the study variables are presented in Table 1. A total of 5,114 twins answered questionnaires on lifestyle-related behaviors during their adolescent years at least once. For 824 twins, epigenetic aging estimates were obtained.

**Table 1.**
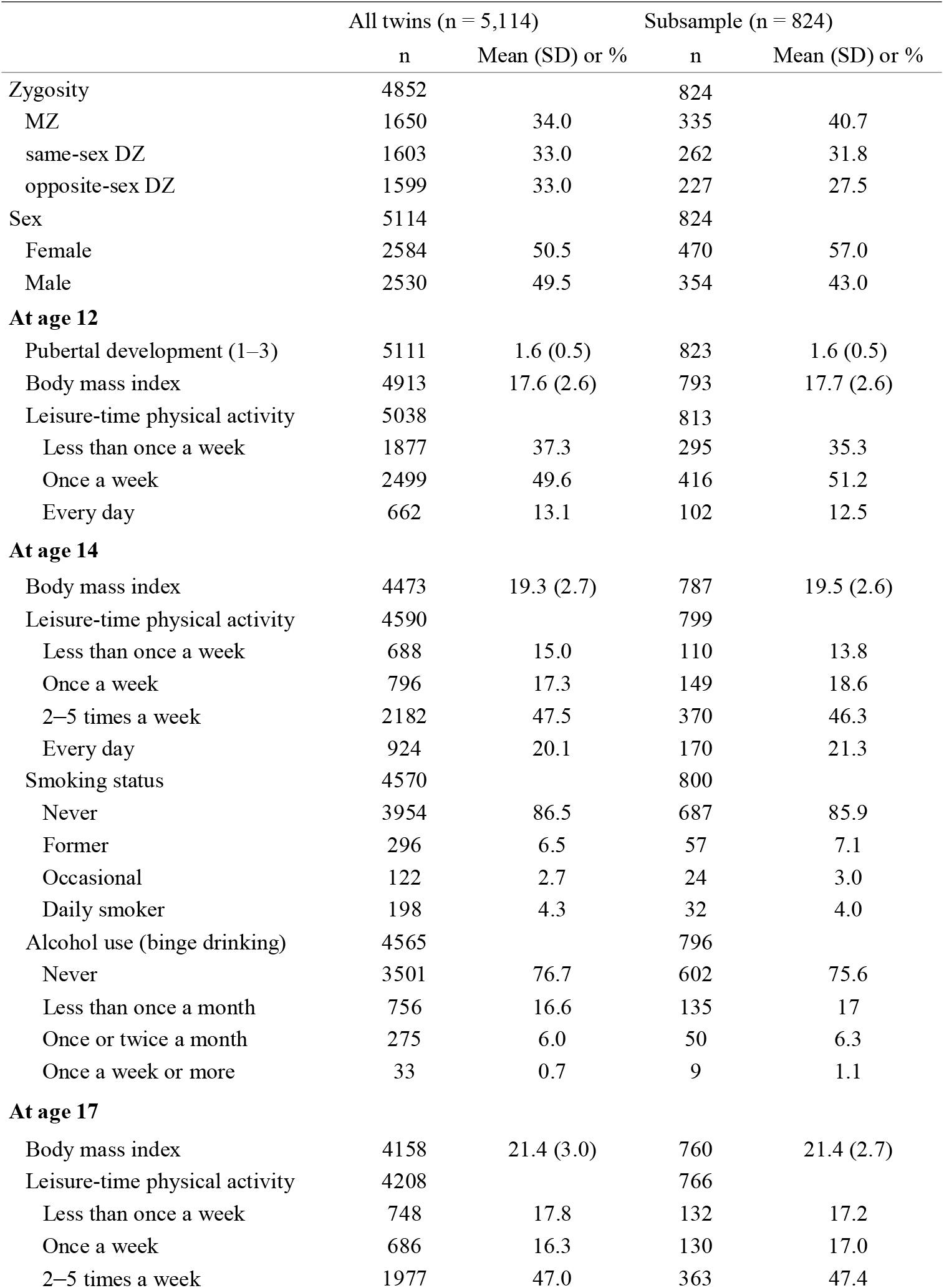
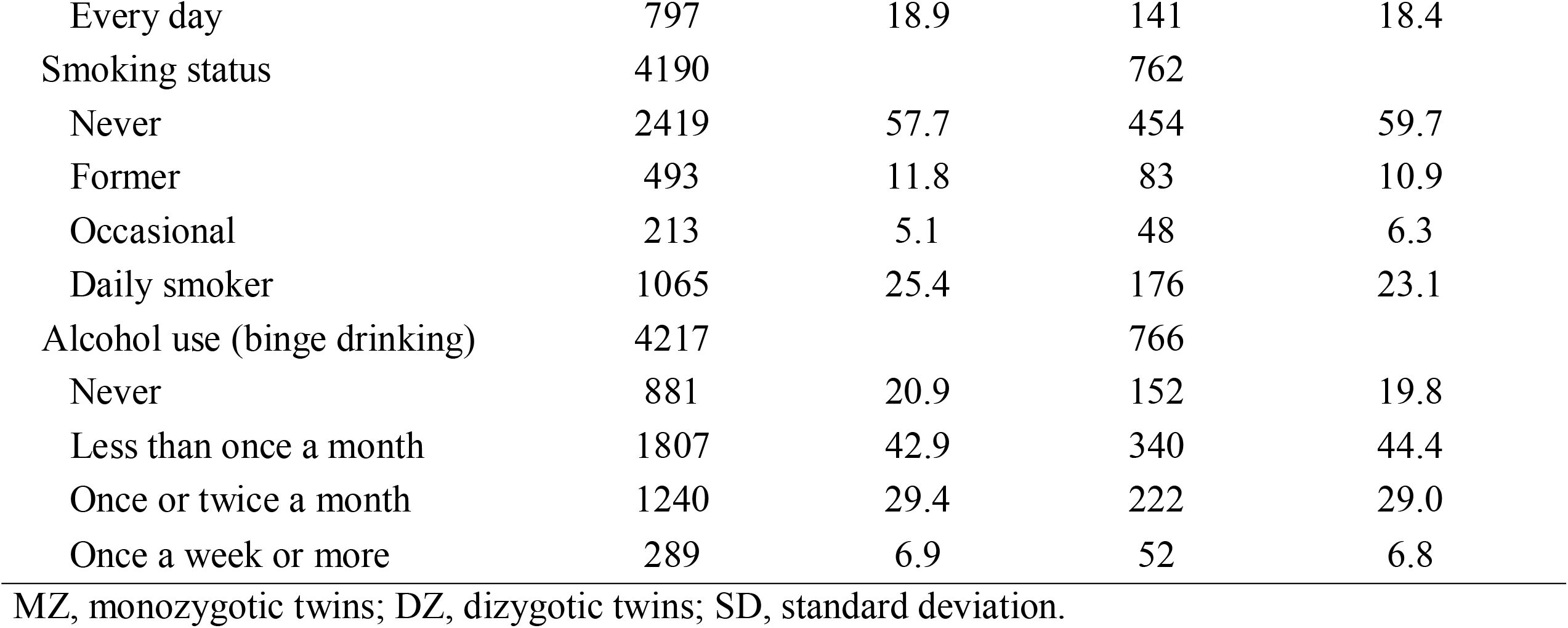
Descriptive statistics of the adolescent lifestyle-related variables in all twins and in the subsample of twins with information on biological aging.

The mean age (SD) of the twins having information on biological aging was 22.4 (0.7) years. The means of the epigenetic age estimates were estimated as follows: Horvath’s clock 28.9 (3.6), Hannum’s clock 18.2 (3.3), DNAm PhenoAge 13.0 (5.3) and DNAm GrimAge 25.2 (3.3) years. The intraclass correlation coefficients (ICCs) of epigenetic aging measures were consistently higher in MZ twin pairs than in DZ twin pairs (Table 2). This suggests an underlying genetic component in biological aging. The correlations between the different epigenetic aging measures ranged from -0.12 to 0.73. The lowest correlation was observed between AA_Horvath_ and DunedinPoAm and between AA_Horvath_ and DunedinPACE. All other correlations were positive. The highest correlations (>0.5) were observed between AA_Hannum_ and AA_Pheno_, AA_Grim_ and DunedinPoAm and DunedinPoAm and DunedinPACE.

**Table 2.** The intraclass correlation coefficients (ICCs) of epigenetic aging measures by zygosity and correlation coefficients between the measures (n = 824)

|  | ICCs (95% CI)<sup>a</sup> |  | Correlation coefficients (95% CI)<sup>a</sup> off-diagonal and means (standard deviations) on the diagonal |  |  |  |  |  |
| --- | --- | --- | --- | --- | --- | --- | --- | --- |
|  | MZ twin pairs | DZ twin pairs | AA<sub>Horvath</sub> | AA<sub>Hannum</sub> | AA<sub>Pheno</sub> | AA<sub>Grim</sub> | DunedinPoAm | DunedinPACE |
| AA<sub>Horvath</sub> | 0.71 (0.63, 0.79) | 0.40 (0.24, 0.55) | 0.00 (3.51) |  |  |  |  |  |
| AA<sub>Hannum</sub> | 0.66 (0.56, 0.76) | 0.32 (0.16, 0.48) | 0.40 (0.33, 0.48) | 0.00 (3.27) |  |  |  |  |
| AA<sub>Pheno</sub> | 0.69 (0.60, 0.78) | 0.16 (0.00, 0.33) | 0.36 (0.29, 0.44) | 0.61 (0.56, 0.66) | 0.00 (5.25) |  |  |  |
| AA<sub>Grim</sub> | 0.72 (0.63, 0.80) | 0.35 (0.15, 0.55) | 0.08 (0.01, 0.16) | 0.32 (0.24, 0.40) | 0.39 (0.33, 0.46) | 0.00 (3.24) |  |  |
| DunedinPoAm | 0.62 (0.52, 0.71) | 0.42 (0.24, 0.60) | -0.05 (-0.12, 0.03) | 0.20 (0.13, 0.27) | 0.41 (0.35, 0.47) | 0.57 (0.52, 0.63) | 1.00 (0.07) |  |
| DunedinPACE | 0.71 (0.64, 0.78) | 0.46 (0.31, 0.61) | -0.04 (-0.11, 0.04) | 0.32 (0.23, 0.40) | 0.49 (0.43, 0.55) | 0.56 (0.50, 0.63) | 0.61 (0.56, 0.67) | 0.88 (0.10) |

CI, confidence interval; AA, age acceleration; MZ, monozygotic; DZ, dizygotic. CIs were corrected for nested sampling.

### Patterns of lifestyle behaviors

Increasing the number of classes continued to improve AIC, BIC and ABIC (Table 3). However, the VLMR and LMR tests indicated that even a solution with four classes would be sufficient. In the fifth step, a class of participants with high BMI was extracted. Previous studies have shown the role of being overweight or obese in biological aging (Lundgren et al., 2021). After including the sixth class, the information criteria still showed considerable improvement, but the AvePPs for several classes were below 0.8. For these reasons, and to have adequate statistical power for subsequent analyses, a five-class solution was considered optimal. The AvePPs ranged from 0.78 to 0.91 for the five-class solution, indicating reasonable classification quality.

**Table 3.** Model fit of the latent class models (n = 5,114).

|  | AIC | BIC | ABIC | VLMR | LMR | Class sizes | AvePP |
| --- | --- | --- | --- | --- | --- | --- | --- |
| 1 | 128842 | 129012 | 128929 |  |  |  |  |
| 2 | 122533 | 122880 | 122711 | <0.001 | <0.001 | 74.0%, 26.0% | 0.95, 0.92 |
| 3 | 119937 | 120460 | 120206 | <0.001 | <0.001 | 44.9%, 40.5%, 14.6% | 0.88, 0.89, 0.93 |
| 4 | 118030 | 118729 | 118389 | <0.001 | <0.001 | 36.4%, 32.7%, 16.7%, 14.2% | 0.83, 0.86, 0.87, 0.92 |
| 5 | 117167 | 118043 | 117617 | 0.529 | 0.530 | 32.0%, 22.8%, 19.9%, 15.9%, 9.5% | 0.78, 0.82, 0.85, 0.88, 0.91 |
| 6 | 116526 | 117578 | 117076 | 0.169 | 0.170 | 31.5%, 18.5%, 15.7%, 14.0%, 12.7%, 7.7% | 0.77, 0.84, 0.83, 0.78, 0.78, 0.90 |
| 7 | 116099 | 117328 | 116731 | 0.043 | 0.044 | 21.0%, 17.5%, 15.2%, 13.8%, 12.9%, 12.8%, 6.9% | 0.73, 0.82, 0.70, 0.77, 0.83, 0.83, 0.91 |
| 8 | 115695 | 117101 | 116418 | 0.407 | 0.408 | 20.3%, 16.2%, 13.6%, 13.5%, 12.3%, 11.3%, 9.3%, 3.4% | 0.72, 0.75, 0.82, 0.71, 0.83, 0.80, 0.82, 0.89 |

AIC, Akaike's information criterion; BIC, Bayesian information criterion; ABIC, sample size-adjusted Bayesian information criterion; VLMR, Vuong–Lo–Mendell–Rubin likelihood ratio test; LMR, Lo–Mendell–Rubin-adjusted likelihood ratio test; AvePP, average posterior probabilities for most likely latent class membership.

Of the participants, 32% fell into the class of healthiest lifestyle habits (C1) (see Figure 2, and the distributions of indicator variables according to the adolescent lifestyle behavior patterns in Supplemental Table S1). They had normal weight, on average, and were more likely to engage in regular LTPA compared to the other groups; most of them were non-smokers and did not use alcohol regularly. Every fifth (19.9%) participant belonged to the second class (C2), characterized by the low mean level of BMI in the range of normal weight for children (low-normal BMI) (Cole et al., 2007). They also had healthy lifestyle habits, but they were not as physically active as the participants in class C1. The participants placed in the third class (C3, 22.8%) had lifestyle habits similar to those of the participants in class C1; however, they had a higher level of BMI in the range of normal weight for children (high-normal BMI). About every tenth (9.5%) of the participants belonged to the fourth class (C4), with the highest level of BMI (high BMI). At each measurement point, the mean BMI level exceeded the cut-off points for overweight in children (Cole et al., 2000). The prevalence of daily smoking was slightly higher in C4 compared to classes C1, C2 and C3. Of the participants, 15.9% were classified in the subgroup characterised by the unhealthiest lifestyle behaviors (C5). Most of them were daily smokers and used alcohol regularly at the age of 17. They also had a lower probability of engaging in regular LTPA compared to the other groups; however, they were of normal weight, on average.

**Figure 2.**
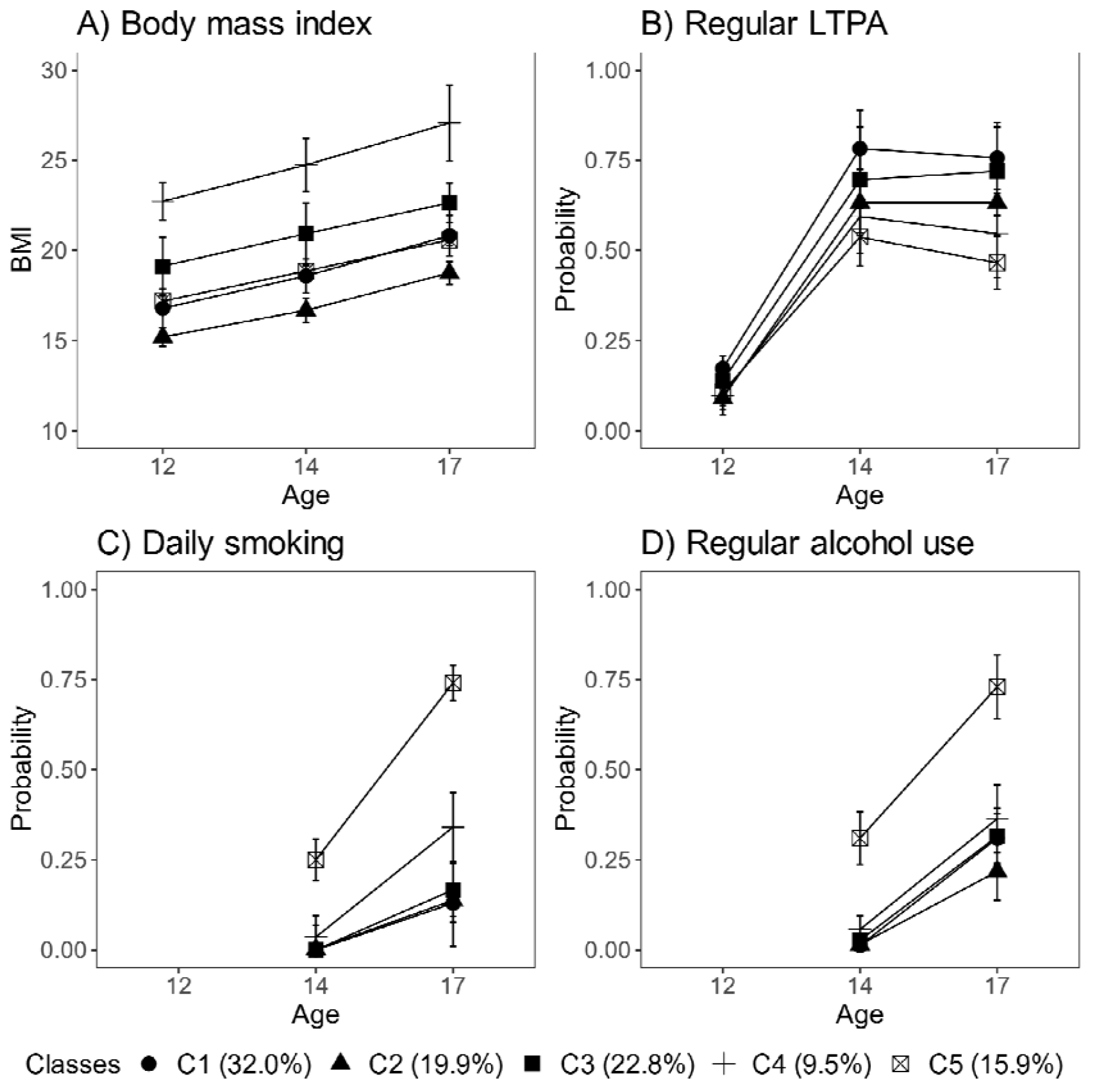
Classes with different lifestyle patterns (n = 5114). Mean and probability profiles (95% confidence intervals) of the indicator variables utilized in the classification: A) body mass index, B) regular LTPA (several times a week), C) daily smoking and D) regular alcohol use (once a month or more). For categorical variables, the probabilities of belonging to the highest categories are presented.

Boys were slightly over-represented in the classes that were most physically active (C1, C3) and had the highest levels of BMI (C3, C4) (percentage of boys: C1: 57.2%, C3: 51.5% and C4: 52.7%), and under-represented in the classes with lowest levels of BMI (C2) and the unhealthiest lifestyle behavior pattern (C5) (C2: 42.7% and C5: 44.1%). There were also differences in pubertal development at baseline between the groups. The subgroups with the highest levels of BMI (C3, C4) and the class with unhealthiest lifestyle habits (C5) were, on average, the most advanced in pubertal development (Mean PDS, C3: 1.67 95% CI: [1.63 to 1.71], C4: 1.69 [1.64 to 1.74] and C5: 1.68 [1.63 to 1.72]), while the class with the healthiest lifestyle pattern (C1) and that with the lowest level of BMI (C2) were less advanced in pubertal development (C1: 1.53 [1.50 to 1.56] and C2: 1.44 [1.41 to 1.47]).

### Differences in biological aging

The distribution of lifestyle behavior patterns in the subsample of participants having information on biological aging was very similar to that in the large cohort data (C1: 33.0%, C2: 16.6%, C3: 20.6%, C4: 10.1%, C5: 19.7%).

There were differences among the classes in AA_Pheno_ (Wald test: *p* = 0.006), AA_Grim_ (*p* = 2.3e-11), DunedinPoAm (*p* = 3.1e-9) and DunedinPACE (*p* = 5.5e-7) in the models adjusted for sex, age and baseline pubertal development. There were no differences in biological aging when Horvath’s clock (*p* = 0.550) and Hannum’s clock (*p* = 0.487) were used. The overall results considering AA_Grim_, DunedinPoAm and DunedinPACE were very similar (Figure 3 and Table 4).

**Figure 3.**
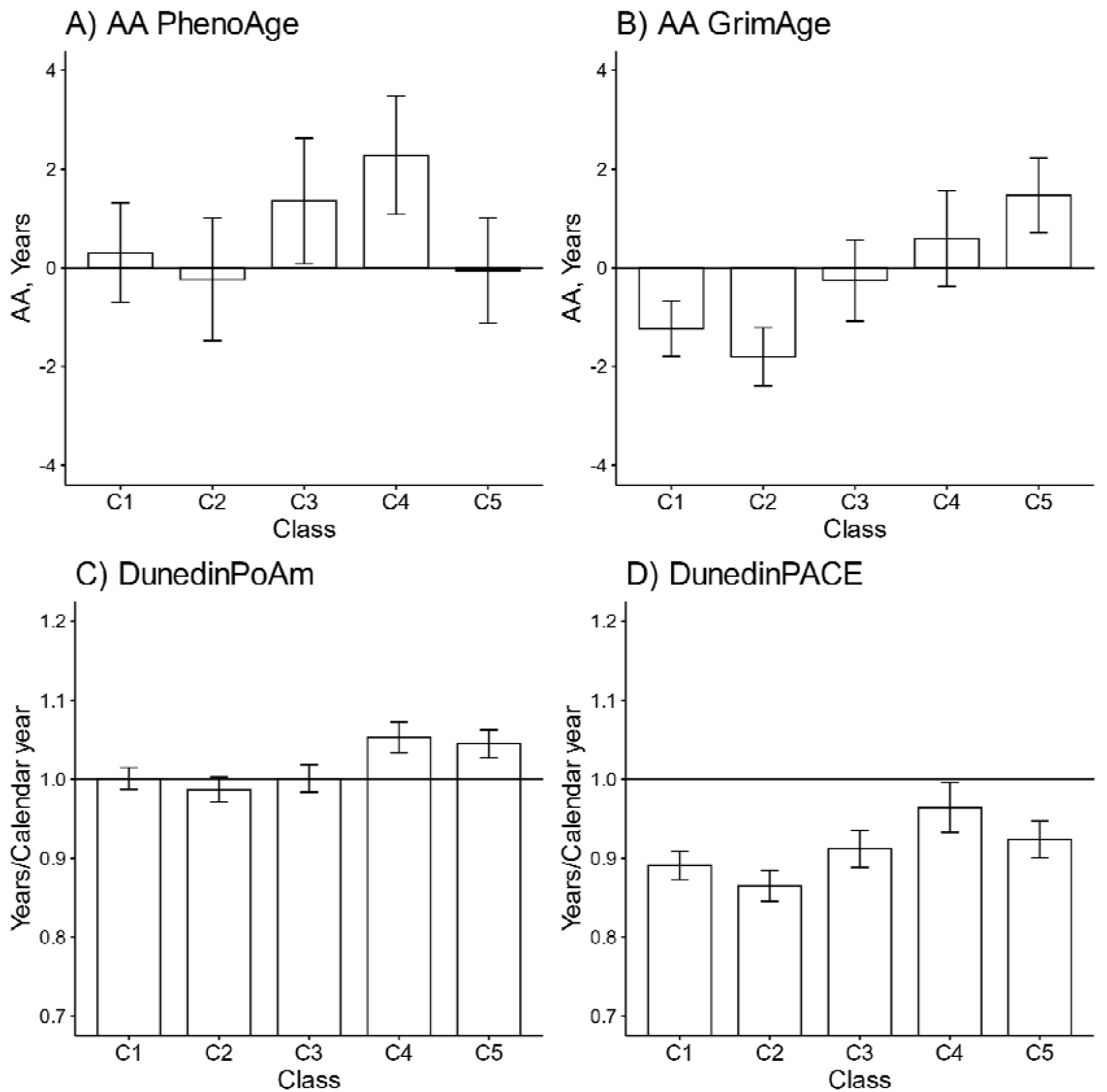
Mean differences between the adolescent lifestyle behavior patterns in biological aging measured with A) DNAm PhenoAge, B) DNAm GrimAge, C) DunedinPoAm and D) DunedinPACE estimators (n = 824). The analysis was adjusted for sex (female), standardized age and baseline pubertal development. Means and 95% confidence intervals are presented. C1 = the class with the healthiest lifestyle pattern, C2 = the class with low–normal BMI, C3 = the class with a healthy lifestyle and high–normal BMI, C4 = the class with high BMI, C5 = the class with the unhealthiest lifestyle pattern. AA, age acceleration.

**Table 4.**
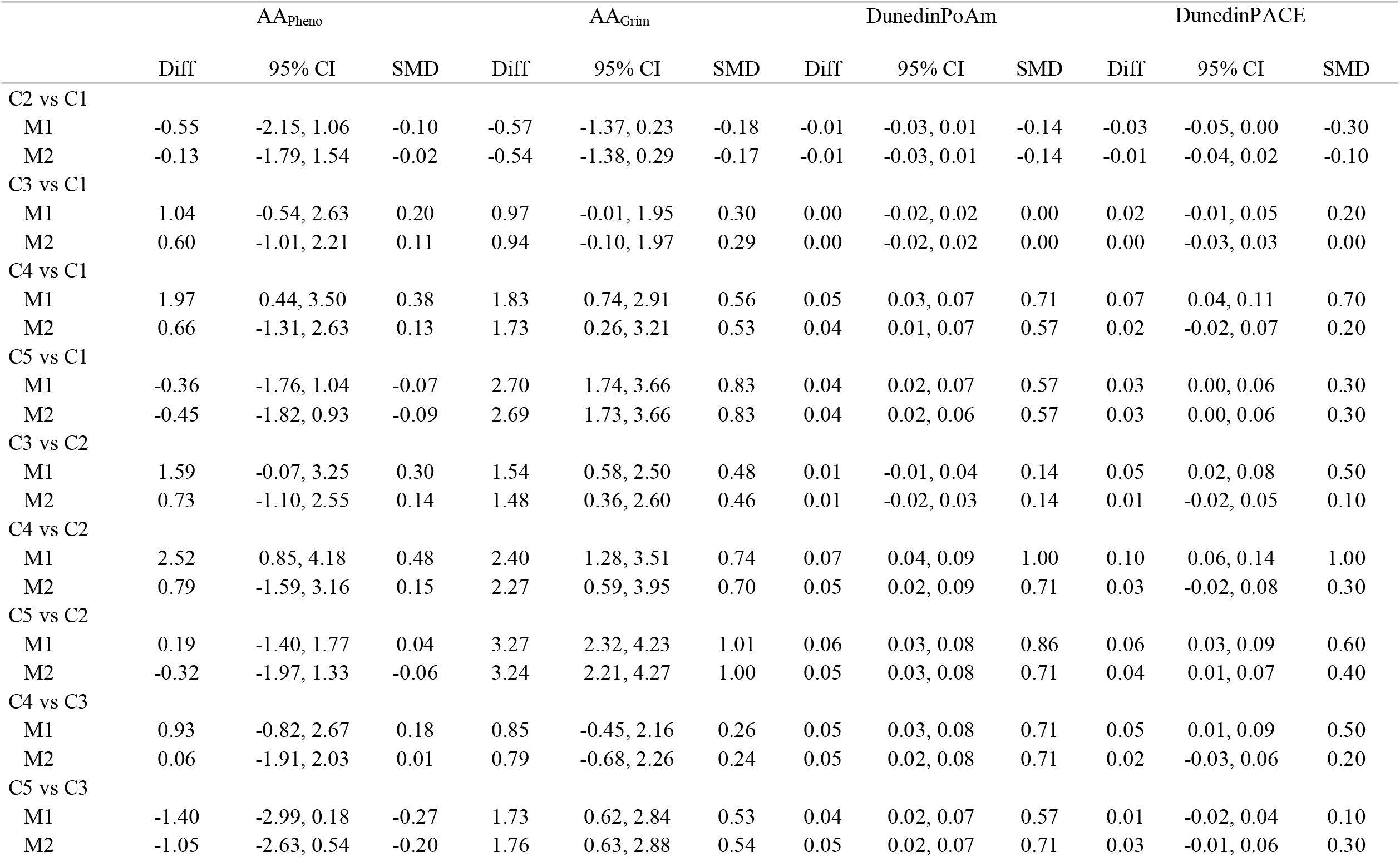
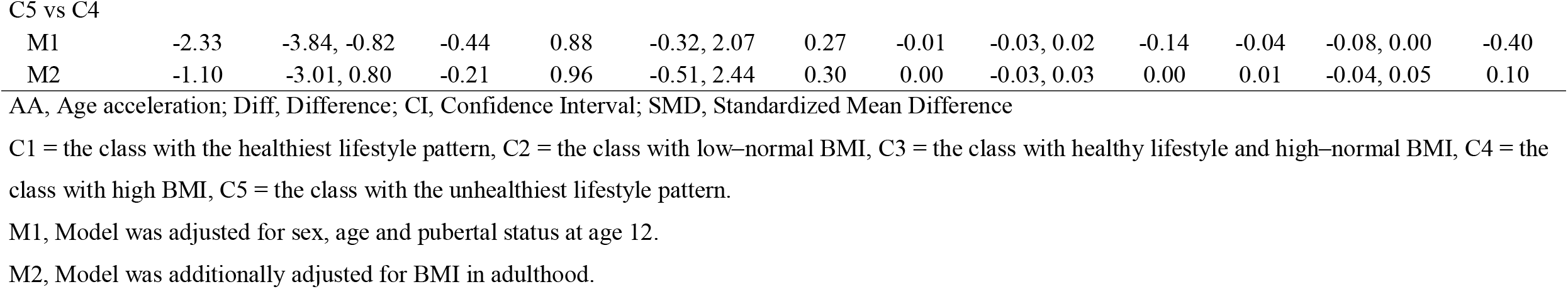
Differences in biological aging between classes with different adolescent lifestyle behavior patterns.

|  | AA<sub>Pheno</sub> |  |  | AA<sub>Grim</sub> |  |  | DunedinPoAm |  |  | DunedinPACE |  |  |
| --- | --- | --- | --- | --- | --- | --- | --- | --- | --- | --- | --- | --- |
|  | Diff | 95% CI | SMD | Diff | 95% CI | SMD | Diff | 95% CI | SMD | Diff | 95% CI | SMD |
| C2 vs C1 |  |  |  |  |  |  |  |  |  |  |  |  |
| M1 | -0.55 | -2.15, 1.06 | -0.10 | -0.57 | -1.37, 0.23 | -0.18 | -0.01 | -0.03, 0.01 | -0.14 | -0.03 | -0.05, 0.00 | -0.30 |
| M2 | -0.13 | -1.79, 1.54 | -0.02 | -0.54 | -1.38, 0.29 | -0.17 | -0.01 | -0.03, 0.01 | -0.14 | -0.01 | -0.04, 0.02 | -0.10 |
| C3 vs C1 |  |  |  |  |  |  |  |  |  |  |  |  |
| M1 | 1.04 | -0.54, 2.63 | 0.20 | 0.97 | -0.01, 1.95 | 0.30 | 0.00 | -0.02, 0.02 | 0.00 | 0.02 | -0.01, 0.05 | 0.20 |
| M2 | 0.60 | -1.01, 2.21 | 0.11 | 0.94 | -0.10, 1.97 | 0.29 | 0.00 | -0.02, 0.02 | 0.00 | 0.00 | -0.03, 0.03 | 0.00 |
| C4 vs C1 |  |  |  |  |  |  |  |  |  |  |  |  |
| M1 | 1.97 | 0.44, 3.50 | 0.38 | 1.83 | 0.74, 2.91 | 0.56 | 0.05 | 0.03, 0.07 | 0.71 | 0.07 | 0.04, 0.11 | 0.70 |
| M2 | 0.66 | -1.31, 2.63 | 0.13 | 1.73 | 0.26, 3.21 | 0.53 | 0.04 | 0.01, 0.07 | 0.57 | 0.02 | -0.02, 0.07 | 0.20 |
| C5 vs C1 |  |  |  |  |  |  |  |  |  |  |  |  |
| M1 | -0.36 | -1.76, 1.04 | -0.07 | 2.70 | 1.74, 3.66 | 0.83 | 0.04 | 0.02, 0.07 | 0.57 | 0.03 | 0.00, 0.06 | 0.30 |
| M2 | -0.45 | -1.82, 0.93 | -0.09 | 2.69 | 1.73, 3.66 | 0.83 | 0.04 | 0.02, 0.06 | 0.57 | 0.03 | 0.00, 0.06 | 0.30 |
| C3 vs C2 |  |  |  |  |  |  |  |  |  |  |  |  |
| M1 | 1.59 | -0.07, 3.25 | 0.30 | 1.54 | 0.58, 2.50 | 0.48 | 0.01 | -0.01, 0.04 | 0.14 | 0.05 | 0.02, 0.08 | 0.50 |
| M2 | 0.73 | -1.10, 2.55 | 0.14 | 1.48 | 0.36, 2.60 | 0.46 | 0.01 | -0.02, 0.03 | 0.14 | 0.01 | -0.02, 0.05 | 0.10 |
| C4 vs C2 |  |  |  |  |  |  |  |  |  |  |  |  |
| M1 | 2.52 | 0.85, 4.18 | 0.48 | 2.40 | 1.28, 3.51 | 0.74 | 0.07 | 0.04, 0.09 | 1.00 | 0.10 | 0.06, 0.14 | 1.00 |
| M2 | 0.79 | -1.59, 3.16 | 0.15 | 2.27 | 0.59, 3.95 | 0.70 | 0.05 | 0.02, 0.09 | 0.71 | 0.03 | -0.02, 0.08 | 0.30 |
| C5 vs C2 |  |  |  |  |  |  |  |  |  |  |  |  |
| M1 | 0.19 | -1.40, 1.77 | 0.04 | 3.27 | 2.32, 4.23 | 1.01 | 0.06 | 0.03, 0.08 | 0.86 | 0.06 | 0.03, 0.09 | 0.60 |
| M2 | -0.32 | -1.97, 1.33 | -0.06 | 3.24 | 2.21, 4.27 | 1.00 | 0.05 | 0.03, 0.08 | 0.71 | 0.04 | 0.01, 0.07 | 0.40 |
| C4 vs C3 |  |  |  |  |  |  |  |  |  |  |  |  |
| M1 | 0.93 | -0.82, 2.67 | 0.18 | 0.85 | -0.45, 2.16 | 0.26 | 0.05 | 0.03, 0.08 | 0.71 | 0.05 | 0.01, 0.09 | 0.50 |
| M2 | 0.06 | -1.91, 2.03 | 0.01 | 0.79 | -0.68, 2.26 | 0.24 | 0.05 | 0.02, 0.08 | 0.71 | 0.02 | -0.03, 0.06 | 0.20 |
| C5 vs C3 |  |  |  |  |  |  |  |  |  |  |  |  |
| M1 | -1.40 | -2.99, 0.18 | -0.27 | 1.73 | 0.62, 2.84 | 0.53 | 0.04 | 0.02, 0.07 | 0.57 | 0.01 | -0.02, 0.04 | 0.10 |
| M2 | -1.05 | -2.63, 0.54 | -0.20 | 1.76 | 0.63, 2.88 | 0.54 | 0.05 | 0.02, 0.07 | 0.71 | 0.03 | -0.01, 0.06 | 0.30 |
| C5 vs C4 |  |  |  |  |  |  |  |  |  |  |  |  |
| M1 | -2.33 | -3.84, -0.82 | -0.44 | 0.88 | -0.32, 2.07 | 0.27 | -0.01 | -0.03, 0.02 | -0.14 | -0.04 | -0.08, 0.00 | -0.40 |
| M2 | -1.10 | -3.01, 0.80 | -0.21 | 0.96 | -0.51, 2.44 | 0.30 | 0.00 | -0.03, 0.03 | 0.00 | 0.01 | -0.04, 0.05 | 0.10 |

AA, Age acceleration; Diff, Difference; CI, Confidence Interval; SMD, Standardized Mean Difference
C1 = the class with the healthiest lifestyle pattern, C2 = the class with low-normal BMI, C3 = the class with healthy lifestyle and high-normal BMI, C4 = the class with high BMI, C5 = the class with the unhealthiest lifestyle pattern.
M1, Model was adjusted for sex, age and pubertal status at age 12.
M2, Model was additionally adjusted for BMI in adulthood.

The group with the unhealthiest lifestyle pattern (C5) was, on average, 1.7–3.3 years biologically older than the groups with healthier lifestyle patterns and normal weight (C1-C3) when DNAm GrimAge was used to assess biological aging (Table 4, M1). Moreover, the unhealthiest group had, an average, 2–3 weeks/calendar year faster pace of biological aging, as measured with DunedinPoAm. The differences in DunedinPACE were very similar to those observed in DunedinPoAm, but there was no difference between the unhealthiest class (C5) and the class with a healthy lifestyle and high-normal BMI (C3).

When DNAm GrimAge was used, the group with a high BMI (C4) was, on average, 1.8–2.4 years biologically older than the two groups with healthier lifestyle patterns (C1 and C2) (Table 4, M1). When measured with the DunedinPoAm estimator, the class had, on average, 3–4 weeks/calendar year faster pace of aging, and when measured with the DunedinPACE estimator, it had 4–5 weeks/calendar year faster pace of aging. Moreover, when DunedinPoAm and DunedinPACE were used, the class had approximately 3 weeks/calendar year faster pace of aging compared to the group with healthy lifestyle with normal-high BMI (C3), and when DunedinPACE was used, the class had 2 weeks/calendar year faster pace of aging compared to the group with unhealthiest lifestyle pattern (C5). When DNAm PhenoAge was used to assess biological aging, only the group with a high BMI stood out. The group was biologically 2.0–2.5 years older than the groups with lower mean levels of BMI (C1–C2, C5). Based on the estimation results of the models, baseline pubertal development was associated with advanced biological aging only when Hannum’s clock was used to derive biological AA (standardised regression coefficient B = 0.10 [0.01 to 0.18]).

After additionally adjusting for BMI in adulthood, the differences in AA_Pheno_ and DunedinPACE between the class of participants with high BMI (C4) and those with lower BMI (C1, C2, C5) were attenuated (Table 4, M2). The differences in biological ageing were only slightly attenuated when the DNAm GrimAge and DunedinPoAm estimators were used.

The differences in the DNAm-based plasma proteins and smoking pack-years are presented in the Supplementary text and Figure S1. In brief, the class with the unhealthiest lifestyle habits (C5) differed unfavourably from the other classes only by DNAm smoking pack-years, while the class of participants with high BMI (C4) stood out by several DNAm-based plasma proteins. The differences in the lifestyle-related factors in adulthood between the lifestyle behavior classes defined in adolescence are presented in the Supplementary text and Figure S2. The lifestyle patterns established during adolescence remained into early adulthood.

### Genetic and environmental effects

Twin pairs with biological aging data on both members of the pair were used in the quantitative genetic modelling to estimate the genetic and environmental components of variance for biological aging (n = 154 monozygotic and 212 dizygotic pairs). Genetic factors explained 62%–73% of variation in biological aging depending on the estimator (Supplementary text and Table S2). The rest of the variation was explained by unshared environmental factors.

The proportion of variation in biological aging in early adulthood explained by adolescent lifestyle behavior patterns was 3.7% for AA_Pheno_, 16.8% for AA_Grim_, 15.4% for DunedinPoAm and 11.1% for DunedinPACE (Figure 4). The association between adolescent lifestyle patterns and biological aging in early adulthood was largely explained by shared genetic factors. Depending on the biological aging estimate, only 0%–3.7% of variation in biological aging was explained by (unshared) environmental factors shared with adolescent lifestyle patterns.

**Figure 4.**
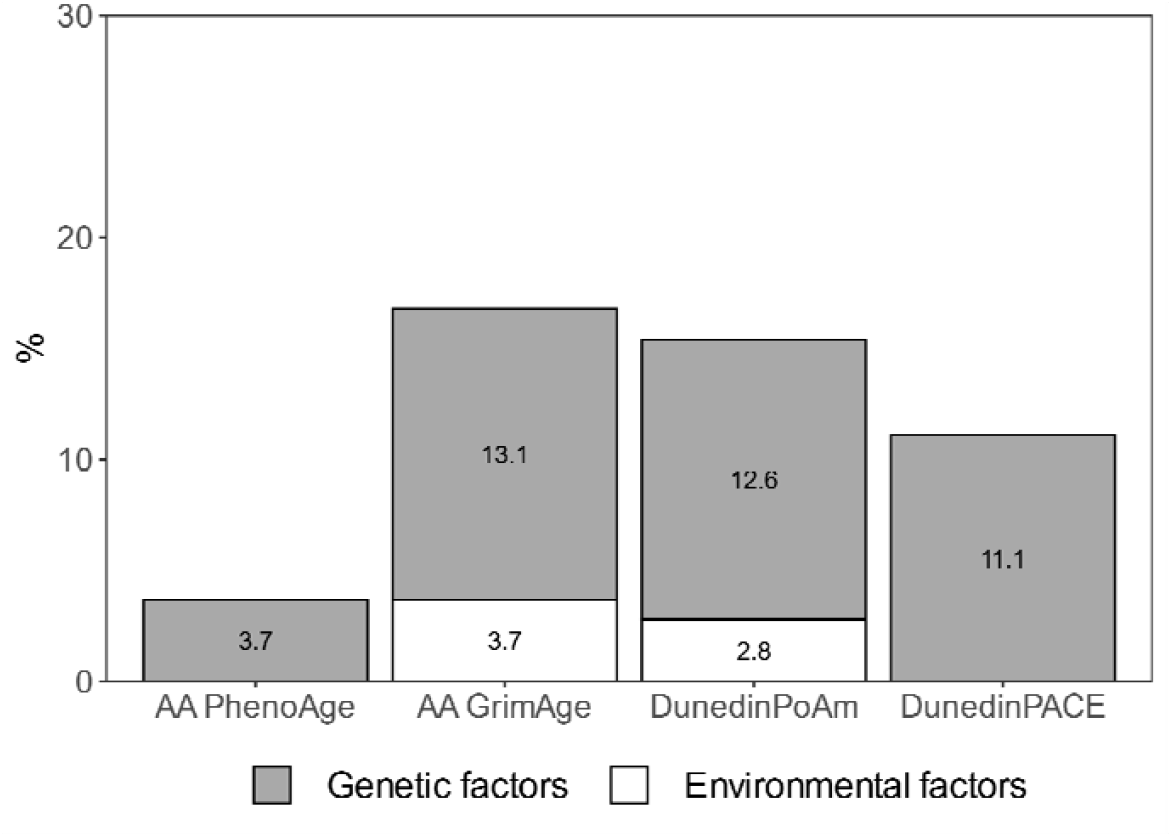
Proportions of variation in biological aging explained by genetic and (unshared) environmental factors shared with adolescent lifestyle patterns. The results are based on the model including additive genetic and non-shared environmental component (AE model). AA, age acceleration.

## DISCUSSION

We conducted a twin study with a longitudinal lifestyle follow-up during the adolescent years and measured biological aging from genome-wide DNAm data using the most recent epigenetic aging clocks. Our findings supported previous studies, which showed that lifestyle-related behaviors tend to cluster in adolescence. In our study, most participants generally followed healthy lifestyle patterns, but we could also identify a group of young adults characterised by higher BMI (10% of all participants) in adolescence, as well as a group (16% of all participants) with more frequent co-occurrence of smoking, binge drinking and low levels of physical activity in adolescence. We observed differences in biological aging between the classes characterised by adolescent lifestyle patterns in young adulthood. Both the class with the overall unhealthiest lifestyle and that with a high BMI were biologically 1.7–3.3 years older (AA_Grim_) than the classes with healthier lifestyle patterns.

Moreover, they had 2–5 weeks/calendar year faster pace of biological aging (DunedinPoAm and DundeinPACE). The differences in lifestyle-related factors were maintained well over the transition from adolescence to young adulthood. However, genetic factors shared with adolescent lifestyle explained most of the observed differences in biological aging. Our findings suggest pleiotropic genetic effects; that is, the same genes affect both adolescent lifestyle and the pace of biological aging.

In our study, when the most recently published epigenetic aging measures were used, the class with the unhealthiest lifestyle was biologically 1.7–3.3 years older (AA_Grim_) and had 2– 3 weeks/calendar year faster pace of biological aging (DunedinPoAm and DundeinPACE) than the classes with healthier patterns. These measures can predict mortality and morbidity, especially cardiometabolic and lung diseases (Belsky et al., 2020, 2022; Lu et al., 2019). A previous meta-analysis showed that the number of healthy lifestyle behaviors is inversely associated with all-cause mortality risk (Loef & Walach, 2012). The mortality risk was up to 66% lower for individuals having multiple healthy behaviors compared to those adhering to an unhealthy lifestyle (smoking, low or high levels of alcohol use, unhealthy diet, no physical activity and overweight). Our results suggested that the accumulation of multiple unhealthy lifestyle habits in adolescence has a more detrimental effect on biological aging than any single lifestyle habit. The unhealthy lifestyle-induced changes in biological aging begin to accumulate in early life and might predispose individuals to premature death in later life.

To the best of our knowledge, this is the first study to investigate common genetic influences underlying lifestyle clusters and biological aging. In our study, the genetic factors shared between adolescent lifestyle and biological aging largely explained the observed differences in biological aging. Our results suggest that individuals who are genetically prone to unhealthy lifestyles or overweight in adolescence are also susceptible to faster biological aging later in young adulthood. These results are supported by McCartney et al. (2021), who showed shared underlying genetic contributions between single lifestyle factors and polygenetic risk scores for epigenetic AA. They concluded that genetic pleiotropy is an underlying mechanism, especially behind associations of BMI and biological aging (AA_Grim_, AA_Pheno_) and smoking and biological aging (AA_Grim_).

To the best of our knowledge, this is also the first study reporting the association between adolescent BMI (relative weight) and biological aging in later life. Previous systematic reviews have concluded that being overweight or obese in childhood and adolescence has a consistent impact on mortality and morbidity in later life (Park et al., 2012; Reilly & Kelly, 2011). In particular, the associations with cardiometabolic morbidity are well-established, but the results of the studies investigating the associations independent of adult BMI are inconclusive (Park et al., 2012). A more recent study showed that early-life body size indirectly predisposes coronary artery disease and type 2 diabetes through body size in adulthood rather than having a direct effect (Richardson et al., 2020). Our results considering biological aging are in line with the existing literature but depend on the epigenetic clock utilized. In our study, the participants assigned to the class that was, on average, overweight in adolescence were biologically older (based on AA_Pheno_, AA_Grim_, DunedinPoAm and DunedinPACE) in young adulthood compared to the classes of normal weight and healthy lifestyle habits. The group stood out, especially when AA_Pheno_ and DunedinPACE were used to measure biological aging, but adult BMI explained the observed differences in these measures. Practically all variance of AA_Pheno_ and DunedinPACE common with adolescent lifestyle was explained by shared genetic factors. Therefore, these measures probably capture aspects of biological aging that are attributed to genetic factors shared with BMI. The differences in AA_Grim_ and DunedinPoAm did not attenuate after additionally controlling for adult BMI, suggesting that higher BMI in adolescence has a direct long-term effect on biological aging measured with these epigenetic clocks.

LTPA is associated with a lower risk of mortality and cardiovascular diseases (Li et al., 2013; Löllgen et al., 2009). Twin studies and genetically informed studies have suggested that genetic pleiotropy can partly explain these frequently observed associations (Karvinen et al., 2015; Sillanpää et al., 2022). Previous studies have shown that LTPA is also associated with slower biological aging (Kankaanpää et al., 2020). In the present study, lower levels of physical activity in adolescence were closely intertwined with other unhealthy behaviors. To fully understand the role of adolescence physical activity in later biological aging would require a more comprehensive analysis of activity patterns, intensities and modes, as well as subgroup analyses that account for other lifestyle factors, such as diet.

Adolescent smoking behavior and alcohol use appeared to be strongly clustered, in line with the findings of a recent systematic review (Whitaker et al., 2021). For this reason, the associations of smoking and alcohol use with biological aging might be difficult to disentangle. Smoking is the most detrimental lifestyle factor, and its’ association with accelerated biological aging has been frequently reported (Oblak et al., 2021). However, the results obtained for the association between alcohol use and biological aging remain unclear (Oblak et al., 2021). A recent study showed that smoking has a causal effect on AA_Grim_, whereas alcohol use did not exhibit such effect (McCartney et al., 2021). Epigenetic methylation changes due to alcohol seem to be much fewer in number and magnitude compared to smoking exposure (Stephenson et al., 2021). In our study, the unhealthiest lifestyle class, in which smoking and alcohol use co-occurred, exhibited accelerated biological aging, especially when GrimAge and DunedinPoAm were used. These epigenetic aging measures are highly sensitive to tobacco exposure (Belsky et al., 2020; Lu et al., 2019). DNAm GrimAge is a composite biomarker comprising seven DNAm surrogates for plasma markers and smoking pack-years, which can predict the time to death (Lu et al., 2019).

DunedinPoAm utilizes a specific CpG site (located within the gene AHRR), the methylation of which is strongly affected by tobacco exposure (Belsky et al., 2020). For these reasons, most of the variation in biological aging, which is explained by environmental factors shared with adolescent lifestyle, is probably due to smoking exposure.

To better understand the observed differences in biological aging, we also studied differences in DNAm-based surrogates included in the DNAm GrimAge estimator (Supplementary text and Figure S1). The class with the unhealthiest lifestyle pattern differed unfavourably from those with healthier habits only in DNAm-based smoking pack-years. The class with a high BMI had increased levels of several DNAm-based plasma markers, including DNAm PAI-1 an TIMP-1, which are associated with markers of inflammation and metabolic conditions (Lu et al., 2019). These findings support the suggestions that AA_Grim_ is a useful biomarker for cardiovascular health and a potential predictor of cardiovascular disease already in young adulthood (Joyce et al., 2021).

Recent studies have yielded inconsistent results regarding the association between pubertal timing and biological aging (Hamlat et al., 2021; Maddock et al., 2021). In our models studying the differences in biological aging across adolescent lifestyle patterns, pubertal development at the age of 12 was not associated with accelerated biological aging in young adulthood (except for AA_Hannum_). Moreover, the class with a high BMI included participants with advanced pubertal development, which might reflect the common genetic background underlying BMI and age at menarche (Kaprio et al., 1995). All these findings support the studies showing that childhood obesity, which tracks forward into adulthood, explains the observed associations between advanced pubertal status and worse cardiovascular health (Bell et al., 2018) and can further reflect the genetic architecture underlying BMI, pubertal development and worse health (Day et al., 2015).

Our study has the following major strengths. Adolescent lifestyle-related patterns were identified using population-based large cohort data (N∼5000), with longitudinal measurements of lifestyle-related factors assessed using validated questionnaires. Moreover, adolescent lifestyle behavior patterns were identified using data-driven LCA. This approach enabled us to use all available data on adolescent lifestyle-related behaviors and to identify the patterns without using artificial cut-off points for the variables. The reciprocal associations between different lifestyle-related factors, as well as their joint association with biological aging, are complex, and individual associations are difficult to interpret. However, our approach produced results with easy interpretation. The data were prospective, and biological aging was assessed with novel epigenetic aging measures, including a recently published DunedinPACE estimator. Furthermore, for the first time, we could evaluate the proportions of genetic and environmental influences underlying adolescent lifestyle as a whole in relation to biological aging by using quantitative genetic modelling. However, our study also has some limitations. Adolescent lifestyle-related behaviors were self-reported and, therefore, might be susceptible to recall bias and bias through social desirability.

In conclusion, later biological aging reflects adolescent lifestyle behavior. Our findings advance research on biological aging by showing that a shared genetic background can underlie both adolescent lifestyle and biological aging measured with epigenetic clocks.

## Data Availability

The DNA methylation age estimates and phenotypes of the subsample are located in the Biobank of the National Institute for Health and Welfare, Finland. All the biobanked data are publicly available for use by qualified researchers following a standardised application procedure. Because of the consent given by study participants and the high degree of identifiability of the twin siblings in Finland, the FTC data cannot be made publicly available. The data are available through the Institute for Molecular Medicine Finland (FIMM) Data Access Committee (DAC) for authorized researchers who have IRB/ethics approval and an institutionally approved study plan. For more details, please contact the FIMM DAC.

## REFERENCES

Asparouhov, T., & Muthén, B. (2021). Auxiliary Variables in Mixture Modeling: Using the BCH Method in Mplus to Estimate a Distal Outcome Model and an Arbitrary Second Model. Mplus Web Notes, 21, 1–27.

Baecke, J. A. H., Burema, J., & Fritjters, J. E. R. (1982). A short questionnaire for the measurement of habitual physical activity in epidemiological studies. Am J Clin Nutr., 96(5), 932–942. https://doi.org/10.1093/ajcn/36.5.936

Bell, J. A., Carslake, D., Wade, K. H., Richmond, R. C., Langdon, R. J., Vincent, E. E., Holmes, M. V., Timpson, N. J., & Davey Smith, G. (2018). Influence of puberty timing on adiposity and cardiometabolic traits: A Mendelian randomisation study. PLoS Medicine, 15(8), 1–25. https://doi.org/10.1371/journal.pmed.1002641

Belsky, D. W., Caspi, A., Arseneault, L., Baccarelli, A., Corcoran, D., Gao, X., Hannon, E., Harrington, H. L., Rasmussen, L. J. H., Houts, R., Huffman, K., Kraus, W. E., Kwon, D., Mill, J., Pieper, C. F., Prinz, J., Poulton, R., Schwartz, J., Sugden, K., … Moffitt, T. E. (2020). Quantification of the pace of biological aging in humans through a blood test, the DunedinPoAm DNA methylation algorithm. ELife, 9, e54870. https://doi.org/10.7554/eLife.54870

Belsky, D. W., Caspi, A., Corcoran, D. L., Sugden, K., Poulton, R., Arseneault, L., Baccarelli, A., Chamarti, K., Gao, X., Hannon, E., Harrington, H. L., Houts, R., Kothari, M., Kwon, D., Mill, J., Schwartz, J., Vokonas, P., Wang, C., Williams, B., & Moffitt, T. E. (2022). DunedinPACE, A DNA methylation biomarker of the Pace of Aging. ELife, 11, e73420. https://doi.org/10.7554/eLife.73420

Biro, F. M., & Deardorff, J. (2013). Identifying opportunities for cancer prevention during preadolescence and adBiro, F. M., & Deardorff, J. (2013). Identifying opportunities for cancer prevention during preadolescence and adolescence: Puberty as a window of susceptibility. Journal of Adole. Journal of Adolescent Health, 52(5), S15–S20. https://doi.org/10.1016/j.jadohealth.2012.09.019

Bucholz, K. K., Cadoret, R., Robert Cloninger, C., Dinwiddie, S. H., Hesselbrock, V. M., Nurnberger, J. I., Reich, T., Schmidt, I., & Schuckit, M. A. (1994). A New, Semi-Structured Psychiatric Interview for Use in Genetic Linkage Studies: A Report on the Reliability of the SSAGA. J Stud Alcohol, 55, 149–158.

Cole, T. J., Flegal, K. M., Nicholls, D., & Jackson, A. A. (2007). Body mass index cut offs to define thinness in children and adolescents: International survey. British Medical Journal, 335(7612), 194–197. https://doi.org/10.1136/bmj.39238.399444.55

Day, F. R., Bulik-Sullivan, B., Hinds, D. A., Finucane, H. K., Murabito, J. M., Tung, J. Y., Ong, K. K., & Perry, J. R. B. (2015). Shared genetic aetiology of puberty timing between sexes and with health-related outcomes. Nature Communications, 6, 1–6. https://doi.org/10.1038/ncomms9842

Etzel, L., Hastings, W. J., Hall, M. A., Heim, C., Meaney, J., Noll, J. G., Donnell, K. J. O., Pokhvisneva, I., Rose, E. J., Schreier, H. M. C., Shenk, C., & Shalev, I. (2021). Obesity and accelerated epigenetic aging in a high-risk cohort of children. MedRxiv. https://doi.org/https://doi.org/10.1101/2021.11.03.21265865

Hamlat, E. J., Prather, A. A., Horvath, S., Belsky, J., & Epel, E. S. (2021). Early life adversity, pubertal timing, and epigenetic age acceleration in adulthood. Developmental Psychobiology, December 2020, 1–13. https://doi.org/10.1002/dev.22085

Hannum, G., Guinney, J., Zhao, L., Zhang, L., Hughes, G., Sadda, S. V., Klotzle, B., Bibikova, M., Fan, J. B., Gao, Y., Deconde, R., Chen, M., Rajapakse, I., Friend, S., Ideker, T., & Zhang, K. (2013). Genome-wide Methylation Profiles Reveal Quantitative Views of Human Aging Rates. Molecular Cell, 49(2), 359–367. https://doi.org/10.1016/j.molcel.2012.10.016

Hartman, S., Li, Z., Nettle, D., & Belsky, J. (2017). External-environmental and internal-health early life predictors of adolescent development. Development and Psychopathology, 29(5), 1839–1849.

Horvath, S. (2013). DNA methylation age of human tissues and cell types. Genome Biology, 14, R115. https://doi.org/10.1186/gb-2013-14-10-r115

Kankaanpää, A., Tolvanen, A., Bollepalli, S., Leskinen, T., Kujala, U. M., Kaprio, J., Ollikainen, M., & Sillanpää, E. (2020). Leisure-Time and Occupational Physical Activity Associates Differently with Epigenetic Aging. Medicine & Science in Sports & Exercise, 53(3), 487–495. https://doi.org/10.1249/mss.0000000000002498

Kaprio, J. (2013). The Finnish Twin Cohort Study: an update. Twin Research and Human Genetics, 16(1), 157–162. https://doi.org/10.1017/thg.2012.142

Kaprio, J., Rimpelä, A., Winter, T., & Viken, R. J. (1995). Common Genetic Influences on BMI and Age at Menarche. Human Biology, 67(5), 739–753.

Karvinen, S., Waller, K., Silvennoinen, M., Koch, L. G., Britton, S. L., Kaprio, J., Kainulainen, H., & Kujala, U. M. (2015). Physical activity in adulthood: Genes and mortality. Scientific Reports, 5, 18259. https://doi.org/10.1038/srep18259

Kuh, D., Ben-Shlomo, Y., Lynch, J., Hallqvist, J., & Power, C. (2003). Life Course Epidemiology. Journal of Epidemiology and Community Health, 57, 778–783. https://doi.org/10.1136/jech.57.10.778

Latvala, A., Rose, R. J., Pulkkinen, L., Dick, D. M., Korhonen, T., & Kaprio, J. (2014). Drinking, smoking, and educational achievement: Cross-lagged associations from adolescence to adulthood. Drug and Alcohol Dependence, 137(1), 106–113. https://doi.org/10.1016/j.drugalcdep.2014.01.016

Levine, M. E., Lu, A. T., Quach, A., Chen, B. H., Assimes, T. L., Bandinelli, S., Hou, L., Baccarelli, A. A., Stewart, J. D., Li, Y., Whitsel, E. A., Wilson, J. G., Reiner1, A. P., Aviv1, A., Lohman, K., Liu, Y., Ferrucci, L., & Horvath, S. (2018). An epigenetic biomarker of aging for lifespan and healthspan. Aging, 10(4), 573–591. https://doi.org/10.18632/aging.101414

Li, J., Loerbroks, A., & Angerer, P. (2013). Physical activity and risk of cardiovascular disease: What does the new epidemiological evidence show? Current Opinion in Cardiology, 28(5), 575–583. https://doi.org/10.1097/HCO.0b013e328364289c

Li, W., Liu, Q., Deng, X., Chen, Y., Liu, S., & Story, M. (2017). Association between obesity and puberty timing: A systematic review and meta-analysis. International Journal of Environmental Research and Public Health, 14(10), 575–583. https://doi.org/10.3390/ijerph14101266

Loef, M., & Walach, H. (2012). The combined effects of healthy lifestyle behaviors on all cause mortality: A systematic review and meta-analysis. Preventive Medicine, 55(3), 163–170. https://doi.org/10.1016/j.ypmed.2012.06.017

Löllgen, H., Böckenhoff, A., & Knapp, G. (2009). Physical activity and all-cause mortality: An updated meta-analysis with different intensity categories. International Journal of Sports Medicine, 30(3), 213–224. https://doi.org/10.1055/s-0028-1128150

López-Otín, C., Blasco, M. A., Partridge, L., Serrano, M., & Kroemer, G. (2013). The Hallmarks of Aging. Cell, 153(6), 1194–1217. https://doi.org/10.1016/j.cell.2013.05.039

Lopez, A. D., Mathers, C. D., Ezzati, M., Jamison, D. T., & Murray, C. J. (2006). Global and regional burden of disease and risk factors, 2001: systematic analysis of population health data. Lancet, 367(9524), 1747–1757. https://doi.org/10.1016/S0140-6736(06)68770-9

Lu, A. T., Quach, A., Wilson, J. G., Reiner, A. P., Aviv, A., Raj, K., Hou, L., Baccarelli, A. A., Li, Y., Stewart, J. D., Whitsel, E. A., Assimes, T. L., Ferrucci, L., & Horvath, S. (2019). DNA methylation GrimAge strongly predicts lifespan and healthspan. Aging, 11(2), 303–327. https://doi.org/10.18632/aging.101684

Lundgren, S., Kuitunen, S., Pietiläinen, K. H., Hurme, M., Kähönen, M., Männistö, S., Perola, M., Lehtimäki, T., Raitakari, O., Kaprio, J., & Ollikainen, M. (2021). BMI is positively associated with accelerated epigenetic aging in twin pairs discordant for BMI. MedRxiv. https://doi.org/10.1101/2021.03.11.21253271

Maddock, J., Castillo-Fernandez, J., Wong, A., Ploubidis, G. B., Kuh, D., Bell, J. T., & Hardy, R. (2021). Childhood growth and development and DNA methylation age in mid-life. Clinical Epigenetics, 13(1), 1–13. https://doi.org/10.1186/s13148-021-01138-x

Maggs, J. L., & Schulenberg, J. E. (2005). Trajectories of Alcohol Use during the Transition to Adulthood. Alcohol Research and Health, 28(4), 195–201.

McCartney, D., Min, J., Richmond, R., Lu, A., Sobczyk, M., Davies, G., & et al. (2021). Genome-wide association studies identify 137 loci for DNA methylation biomarkers of ageing. Genome Biology, 22(194), 1–25. https://doi.org/10.1186/s13059-021-02398-9

Muthén, L. K., & Muthén, B. O. (1998-2018). Mplus User’s Guide. Eight Edition. Los Angeles, CA: Muthén & Muthén. Available from: https://www.statmodel.com

Neale, M. C., & Cardon, L. R. (1992). Methodology for Genetic Studies of Twins and Families. Dordrecht, the Netherlands: Kluver Academic Publisher.

Oblak, L., van der Zaag, J., Higgins-Chen, A. T., Levine, M. E., & Boks, M. P. (2021). A systematic review of biological, social and environmental factors associated with epigenetic clock acceleration. Ageing Research Reviews, 69, 101348. https://doi.org/10.1016/j.arr.2021.101348

Osmond, C., & Barker, D. J. P. (2000). Fetal, Infant, and Childhood Growth Are Predictors of Coronary Heart Disease, Diabetes, and Hypertension in Adult Men and Women. Environmental Health Perspectives, 108, 545–553.

Park, M. H., Falconer, C., Viner, R. M., & Kinra, S. (2012). The impact of childhood obesity on morbidity and mortality in adulthood: A systematic review. Obesity Reviews, 13(11), 985–1000. https://doi.org/10.1111/j.1467-789X.2012.01015.x

Petersen, A. C., Crockett, L., Richards, M., & Boxer, A. (1988). A self-report measure of pubertal status: Reliability, validity, and initial norms. Journal of Youth and Adosecence, 17, 117–133.

Power, C., Kuh, D., & Morton, S. (2013). From Developmental Origins of Adult Disease to Life Course Research on Adult Disease and Aging: Insights from Birth Cohort Studies. The Annual Review of Public Health, 34, 7–28. https://doi.org/10.1146/annurev-publhealth-031912-114423

Prentice, P., & Viner, R. M. (2013). Pubertal timing and adult obesity and cardiometabolic risk in women and men: A systematic review and meta-analysis. International Journal of Obesity, 37(8), 1036–1043. https://doi.org/10.1038/ijo.2012.177

Raffington, L., Belsky, D. W., Kothari, M., Malanchini, M., Tucker-Drob, E. M., & Harden, K. P. (2021). Socioeconomic disadvantage and the pace of biological aging in children. Pediatrics, 147(6), e2020024406. https://doi.org/10.1542/peds.2020-024406

Reilly, J. J., & Kelly, J. (2011). Long-term impact of overweight and obesity in childhood and adolescence on morbidity and premature mortality in adulthood: Systematic review. International Journal of Obesity, 35(7), 891–898. https://doi.org/10.1038/ijo.2010.222

Richardson, M. T., Ainsworth, B. E., Wu, H. C., Jacobs, D. R., & Leon, A. S. (1995). Ability of the atherosclerosis risk in communities (ARIC)/baecke questionnaire to assess leisure-time physical activity. International Journal of Epidemiology, 24(4), 685–693. https://doi.org/10.1093/ije/24.4.685

Richardson, T. G., Sanderson, E., Elsworth, B., Tilling, K., & Smith, G. D. (2020). Use of genetic variation to separate the effects of early and later life adiposity on disease risk: Mendelian randomisation study. The BMJ, 369, 1–12. https://doi.org/10.1136/bmj.m1203

Rose, R. J., Salvatore, J. E., Aaltonen, S., Barr, P. B., Bogl, L. H., Byers, H. A., Heikkilä, K., Korhonen, T., Latvala, A., Palviainen, T., Ranjit, A., Whipp, A. M., Pulkkinen, L., Dick, D. M., & Kaprio, J. (2019). FinnTwin12 Cohort: An Updated Review. Twin Research and Human Genetics, 22(5), 302–311. https://doi.org/10.1017/thg.2019.83

Rovio, S. P., Yang, X., Kankaanpää, A., Aalto, V., Hirvensalo, M., Telama, R., Pahkala, K., Hutri-Kähönen, N., Viikari, J. S. A., Raitakari, O. T., & Tammelin, T. H. (2018). Longitudinal physical activity trajectories from childhood to adulthood and their determinants: The Young Finns Study. Scandinavian Journal of Medicine and Science in Sports, 28(3), 1073–1083. https://doi.org/10.1111/sms.12988

Salin, K., Kankaanpää, A., Hirvensalo, M., Lounassalo, I., Yang, X., Magnussen, C. G., Hutri-Kähönen, N., Rovio, S., Viikari, J., Raitakari, O. T., & Tammelin, T. H. (2019). Smoking and physical activity trajectories from childhood to midlife. International Journal of Environmental Research and Public Health, 16(6). https://doi.org/10.3390/ijerph16060974

Savage, J. E., Rose, R. J., Pulkkinen, L., Silventoinen, K., Korhonen, T., Kaprio, J., Gillespie, N., & DIck, D. M. (2018). Early maturation and substance use across adolescence and young adulthood: A longitudinal study of Finnish twins. Development and Psychopathology, 30(1), 79–92. https://doi.org/10.1017/S0954579417000487

Sillanpää, E., Palviainen, T., Ripatti, S., Kujala, U. M., & Kaprio, J. (2022). Polygenic Score for Physical Activity Is Associated with Multiple Common Diseases. Medicine & Science in Sports & Exercise, 54(2), 280–287. https://doi.org/10.1249/mss.0000000000002788

Simpkin, A. J., Howe, L. D., Tilling, K., Gaunt, T. R., Lyttleton, O., McArdle, W. L., Ring, S. M., Horvath, S., Smith, G. D., & Relton, C. L. (2017). The epigenetic clock and physical development during childhood and adolescence: longitudinal analysis from a UK birth cohort. International Journal of Epidemiology, 46(2), 549–558. https://doi.org/10.1093/ije/dyw307

Stephenson, M., Bollepalli, S., Cazaly, E., Salvatore, J. E., Barr, P., Rose, R. J., Dick, D., Kaprio, J., & Ollikainen, M. (2021). Associations of Alcohol Consumption With EpigenomelWide DNA Methylation and Epigenetic Age Acceleration: Individual-Level and Co-twin Comparison Analyses. Alcohol Clin Exp Res, 45(2), 318–328.

Wehkalampi, K., Silventoinen, K., Kaprio, J., Dick, D. M., Rose, R. J., Pulkkinen, L., & Dunkel, L. (2008). Genetic and environmental influences on pubertal timing assessed by height growth. American Journal of Human Biology, 20(4), 417–423. https://doi.org/10.1002/ajhb.20748

Whitaker, V., Oldham, M., Boyd, J., Fairbrother, H., Curtis, P., Meier, P., & Holmes, J. (2021). Clustering of health-related behaviours within children aged 11–16: a systematic review. BMC Public Health, 21(1), 1–12. https://doi.org/10.1186/s12889-020-10140-6

