## Supplementary material for "The role of adolescent lifestyle habits in biological aging: A prospective twin study"

**SUPPLEMENTARY TEXT**

**Methods**

**DNA methylation and assessment of biological age**

Genomic DNA was extracted from peripheral blood samples using commercial kits. High molecular weight DNA samples (1 μg) were bisulfite converted using EZ-96 DNA methylation-Gold Kit (Zymo Research, Irvine, CA, USA) according to the manufacturer's protocol. The twins and co-twins were randomly distributed across plates, with both twins from a pair on the same plate. DNA methylation (DNAm) profiles were obtained using Illumina’s Infinium HumanMethylation450 BeadChip or the Infinium MethylationEPIC BeadChip (Illumina, San Diego, CA, USA). The Illumina BeadChips measure single-CpG resolution DNAm levels across the human genome. With these assays, it is possible to interrogate over 450,000 (450k) or 850,000 (EPIC) methylation sites quantitatively across the genome at single-nucleotide resolution. Methylation data were preprocessed using R package minfi (Aryee et al., 2014). We calculated detection p values comparing total signal for each probe to the background signal level, to evaluate quality of the samples (Maksimovic et al., 2017). Samples of poor quality (mean detection p > .01) were excluded from further analysis. Data were normalized by using the single-sample Noob normalization method, which is suitable for datasets originating from different platforms (Fortin et al., 2017). Beta values representing CpG methylation levels were calculated as ratio of methylated intensities (M) to the overall intensities (Beta value=M/(M+U+100), where U is unmethylated probe intensity). β values were used to calculate the estimates of epigenetic aging.

**Statistical analysis**

**Univariate modelling**

The variance in the epigenetic aging measures was decomposed into the latent variables representing additive genetic (A), dominant genetic (D) or shared environmental (C) and non-shared environmental (E) components (ACE model or ADE model) (Neale & Cardon, 1992). The sequences of the models were fitted (ACE, ADE, AE, CE and E). Because dominance in the absence of additive effects is rare, the model including D and E components (DE-model) was omitted. We used Satorra-Bentler scaled chi-squared (χ^2^)-test, the comparative fit index (CFI), the Tucker–Lewis index (TLI), the root mean square error of approximation (RMSEA), and the standardised root-mean-square residual (SRMR) to evaluate the goodness-of-fit of the models. The model fits the data well if the χ^2^-test is not statistically significant (P > 0.05), CFI and TLI values are close to 0.95, the RMSEA value is below 0.06, and the SRMR value is below 0.08 (Hu & Bentler, 1999). Moreover, Bayesian information criterion (BIC) was used to compare non-nested models. A lower BIC value indicates a better model fit. The most parsimonious model with the sufficient fit to the data was considered optimal.

**Results**

**Univariate modelling**

The estimation results of the univariate models are presented in Table S2. The model including additive genetic and non-shared environmental component (AE model) was considered optimal for all the epigenetic aging measures. Generally, ACE and ADE fit the data about as well, and models without genetic component (CE model) provided significantly worse fit. Based on these results, AE model was also chosen for the further modelling of the residual term of biological aging.

**Differences in DNAm-based surrogates between the adolescent lifestyle behavior patterns**

Overall, after controlling for sex, age and baseline pubertal development there were differences in DNAm-based ADM (Wald test: *p* = 0.010), B2M (*p* = 0.014) and Packyrs (*p* = 1.3e-5), but not in DNAm-based cystatin C (*p* = 0.140), GDF15 (*p* = 0.228), Leptin (*p* = 0.228), PAI-1 (*p* = 0.055), TIMP-1 (*p* = 0.089) between the adolescent lifestyle behavior patterns. The class with the unhealthiest lifestyle habits (C5) differed unfavorably from the other classes only by DNAm smoking pack-years (Figure S1). The class of participants with high BMI stood out by several DNAm-based plasma proteins including DNAm ADM, PAI-1 and TIMP-1.

**Differences in lifestyle-relted factors in adulthood between the adolescent lifestyle behavior patterns**

The differences in lifestyle-related factors were maintained well over the transition from adolescence to young adulthood (Figure S2). The ranking of the classes by each lifestyle-related factor in young adulthood was practically the same as the ranking based on those measured in adolescence.

**References**

Aryee, M. J., Jaffe, A. E., Corrada-Bravo, H., Ladd-Acosta, C., Feinberg, A. P., Hansen, K. D., & Irizarry, R. A. (2014). Minfi: A flexible and comprehensive Bioconductor package for the analysis of Infinium DNA methylation microarrays. *Bioinformatics*, *30*(10), 1363–1369. https://doi.org/10.1093/bioinformatics/btu049

Fortin, J. P., Triche, T. J., & Hansen, K. D. (2017). Preprocessing, normalization and integration of the Illumina HumanMethylationEPIC array with minfi. *Bioinformatics*, *33*(4), 558–560. https://doi.org/10.1093/bioinformatics/btw691

Hu, L., & Bentler, P. M. (1999). Cutoff criteria for fit indexes in covariance structure analysis: Conventional criteria versus new alternatives. *Structural Equation Modeling: A Multidisciplinary Journal*, *6*(1), 1–55. https://doi.org/10.1080/10705519909540118

Maksimovic, J., Phipson, B., & Oshlack, A. (2017). A cross-package Bioconductor workflow for analysing methylation array data [version 3; peer review: 4 approved]. *F1000Res*, *5*, 1281. https://doi.org/10.12688/f1000research.8839.3

Neale, M. C., & Cardon, L. R. (1992). *Methodology for Genetic Studies of Twins and Families*. Dordrecht, the Netherlands: Kluver Academic Publisher.

.

| Table S1. The classes with different adolescent lifestyle behavior patterns (n = 5114). Mean and probability profiles of the indicator variables utilized in the classification. | | | | | | | | | | |
| --- | --- | --- | --- | --- | --- | --- | --- | --- | --- | --- |
|  | C1 (32.0%) | | C2 (19.9%) | | C3 (22.8%) | | C4 (9.5%) | | C5 (15.9%) | |
|  | Est | 95% CI | Est | 95% CI | Est | 95% CI | Est | 95% CI | Est | 95% CI |
| Body mass index | | | |  |  |  |  |  |  |  |
| At age of 12 years | 16.8 | 15.7, 17.3 | 15.2 | 14.7, 15.5 | 19.1 | 17.5, 20.0 | 22.7 | 21.7, 23.3 | 17.2 | 16.9, 17.3 |
| At age of 14 years | 18.6 | 17.6, 19.1 | 16.7 | 16.0, 17.0 | 20.9 | 19.2, 21.8 | 24.8 | 23.3, 25.5 | 18.9 | 18.6, 19.0 |
| At age of 17 years | 20.8 | 19.7, 21.4 | 18.8 | 18.1, 19.1 | 22.6 | 21.6, 23.2 | 27.1 | 25.0, 28.1 | 20.6 | 20.3, 20.8 |
| Leisure-time physical activity | | |  |  |  |  |  |  |  |  |
| At age of 12 years |  |  |  |  |  |  |  |  |  |  |
| Less than once a week | 0.29 | 0.22, 0.37 | 0.45 | 0.39, 0.51 | 0.35 | 0.26, 0.43 | 0.44 | 0.37, 0.50 | 0.44 | 0.39, 0.48 |
| Once a week | 0.54 | 0.48, 0.59 | 0.46 | 0.41, 0.50 | 0.52 | 0.47, 0.56 | 0.47 | 0.39, 0.54 | 0.46 | 0.41, 0.50 |
| Every day | 0.17 | 0.14, 0.21 | 0.09 | 0.04, 0.14 | 0.14 | 0.07, 0.21 | 0.10 | 0.06, 0.13 | 0.11 | 0.08, 0.14 |
| At age of 14 years |  |  |  |  |  |  |  |  |  |  |
| Less than once a week | 0.08 | 0.05, 0.11 | 0.17 | 0.12, 0.22 | 0.14 | 0.07, 0.22 | 0.18 | 0.13, 0.23 | 0.27 | 0.22, 0.31 |
| Once a week | 0.14 | 0.07, 0.20 | 0.20 | 0.17, 0.24 | 0.16 | 0.10, 0.23 | 0.23 | 0.17, 0.28 | 0.20 | 0.16, 0.23 |
| 2‒5 times a week | 0.52 | 0.45, 0.59 | 0.45 | 0.41, 0.49 | 0.51 | 0.43, 0.59 | 0.43 | 0.37, 0.49 | 0.40 | 0.35, 0.45 |
| Every day | 0.27 | 0.23, 0.30 | 0.18 | 0.13, 0.23 | 0.19 | 0.12, 0.25 | 0.17 | 0.12, 0.21 | 0.14 | 0.10, 0.17 |
| At age of 17 years |  |  |  |  |  |  |  |  |  |  |
| Less than once a week | 0.10 | 0.05, 0.14 | 0.19 | 0.14, 0.23 | 0.13 | 0.06, 0.20 | 0.27 | 0.19, 0.35 | 0.35 | 0.29, 0.40 |
| Once a week | 0.15 | 0.11, 0.18 | 0.18 | 0.15, 0.21 | 0.15 | 0.11, 0.19 | 0.18 | 0.14, 0.23 | 0.19 | 0.15, 0.23 |
| 2‒5 times a week | 0.50 | 0.44, 0.56 | 0.45 | 0.41, 0.49 | 0.53 | 0.48, 0.57 | 0.44 | 0.36, 0.52 | 0.36 | 0.32, 0.41 |
| Every day | 0.26 | 0.22, 0.29 | 0.18 | 0.13, 0.23 | 0.20 | 0.12, 0.27 | 0.11 | 0.07, 0.15 | 0.10 | 0.07, 0.13 |
| Smoking status |  |  |  |  |  |  |  |  |  |  |
| At age of 14 years |  |  |  |  |  |  |  |  |  |  |
| Never | 0.99 | 0.98, 1.00 | 0.98 | 0.95, 1.00 | 0.97 | 0.95, 1.00 | 0.83 | 0.74, 0.93 | 0.33 | 0.24, 0.43 |
| Former | 0.01 | 0.00, 0.02 | 0.02 | 0.00, 0.03 | 0.02 | 0.00, 0.04 | 0.09 | 0.04, 0.14 | 0.29 | 0.24, 0.34 |
| Occasional | 0.00 |  | 0.01 | -0.01, 0.02 | 0.00 | 0.00, 0.01 | 0.04 | 0.01, 0.07 | 0.13 | 0.10, 0.16 |
| Daily smoker | 0.00 |  | 0.00 | 0.00, 0.01 | 0.00 |  | 0.04 | 0.00, 0.07 | 0.25 | 0.19, 0.31 |
| At age of 17 years |  |  |  |  |  |  |  |  |  |  |
| Never | 0.69 | 0.61, 0.77 | 0.73 | 0.65, 0.81 | 0.68 | 0.59, 0.78 | 0.50 | 0.41, 0.59 | 0.03 | 0.00, 0.06 |
| Former | 0.12 | 0.09, 0.15 | 0.09 | 0.05, 0.13 | 0.12 | 0.07, 0.16 | 0.11 | 0.06, 0.16 | 0.15 | 0.12, 0.19 |
| Occasional | 0.06 | 0.04, 0.07 | 0.04 | 0.02, 0.06 | 0.04 | 0.01, 0.06 | 0.05 | 0.02, 0.07 | 0.07 | 0.05, 0.10 |
| Daily smoker | 0.13 | 0.08, 0.18 | 0.14 | 0.09, 0.18 | 0.17 | 0.09, 0.24 | 0.34 | 0.24, 0.44 | 0.74 | 0.69, 0.79 |
| Alcohol use (binge drinking) | | |  |  |  |  |  |  |  |  |
| At age of 14 year |  |  |  |  |  |  |  |  |  |  |
| Never | 0.88 | 0.85, 0.91 | 0.94 | 0.90, 0.97 | 0.84 | 0.79, 0.89 | 0.76 | 0.69, 0.83 | 0.23 | 0.15, 0.31 |
| Less than once a month | 0.11 | 0.08, 0.14 | 0.05 | 0.02, 0.08 | 0.13 | 0.09, 0.17 | 0.18 | 0.12, 0.24 | 0.46 | 0.41, 0.51 |
| Once or twice a month | 0.01 | 0.00, 0.02 | 0.02 | 0.00, 0.03 | 0.03 | 0.01, 0.04 | 0.05 | 0.02, 0.08 | 0.27 | 0.22, 0.32 |
| Once a week or more | 0.00 |  | 0.00 |  | 0.00 |  | 0.00 | 0.00, 0.01 | 0.04 | 0.02, 0.06 |
| At age of 17 years |  |  |  |  |  |  |  |  |  |  |
| Never | 0.21 | 0.18, 0.25 | 0.33 | 0.26, 0.41 | 0.22 | 0.16, 0.28 | 0.23 | 0.15, 0.30 | 0.01 | 0.00, 0.02 |
| Less than once a month | 0.48 | 0.43, 0.52 | 0.45 | 0.40, 0.49 | 0.46 | 0.41, 0.52 | 0.41 | 0.35, 0.47 | 0.26 | 0.22, 0.31 |
| Once or twice a month | 0.28 | 0.24, 0.32 | 0.18 | 0.12, 0.24 | 0.28 | 0.23, 0.33 | 0.29 | 0.23, 0.35 | 0.51 | 0.46, 0.55 |
| Once a week or more | 0.03 | 0.00, 0.06 | 0.04 | 0.02, 0.06 | 0.03 | 0.00, 0.06 | 0.08 | 0.04, 0.11 | 0.22 | 0.18, 0.26 |
| Est, Estimated mean or probability; CI, Confidence Interval; C1 = the class with the healthiest lifestyle pattern, C2 = the class with low-normal BMI, C3 = the class with healthy lifestyle and high-normal BMI, C4 = the class with high BMI, C5 = the class with the unhealthiest lifestyle pattern. | | | | | | | | | | |

| Table S2. The estimation results of the univariate model for biological aging among young adult twin pairs (MZ *n* = 154, DZ *n* = 211) | | | | | | | | | | | | | | | | | |
| --- | --- | --- | --- | --- | --- | --- | --- | --- | --- | --- | --- | --- | --- | --- | --- | --- | --- |
|  | Model fit | | | | | | | | | Parameter estimates and their 95% confidence intervals | | | | | | | |
|  | Χ^2^ | df | SC | *P* | CFI | TLI | RMSEA | SRMR | BIC | a^2^ / total | | c^2^ or d^2^ / total | | e^2^ / total | | total | |
| AA_Pheno_ | | | | | | | | | | | | | | | | | |
| ACE | 5.2 | 3 | 1.27 | 0.155 | 0.98 | 0.99 | 0.06 | 0.06 | 2009 | 0.65 | 0.56, 0.74 | 0.00 |  | 0.35 | 0.26, 0.45 | 1.00 | 0.89, 1.12 |
| ADE | 0.6 | 3 | 0.99 | 0.904 | 1.00 | 1.02 | 0.00 | 0.02 | 2003 | 0.03 | -0.46, 0.51 | 0.65 | 0.15, 1.15 | 0.33 | 0.25, 0.41 | 0.99 | 0.88, 1.09 |
| **AE** | **7.0** | **4** | **0.96** | **0.136** | **0.97** | **0.99** | **0.06** | **0.06** | **2003** | **0.65** | **0.56, 0.74** | **-** |  | **0.35** | **0.26, 0.45** | **1.00** | **0.89, 1.12** |
| CE | 43.5 | 4 | 0.96 | <0.001 | 0.60 | 0.80 | 0.23 | 0.11 | 2038 | - |  | 0.39 | 0.30, 0.48 | 0.61 | 0.52, 0.70 | 0.99 | 0.88, 1.10 |
| E | 107 | 5 | 0.96 | <0.001 | 0.00 | 0.59 | 0.33 | 0.21 | 2093 | - |  | - |  | 1.00 |  | 0.99 | 0.88, 1.10 |
| AA_Grim_ | | | | | | | | | | | | | | | | | |
| ACE | 4.3 | 3 | 2.05 | 0.231 | 0.99 | 0.99 | 0.05 | 0.09 | 1989 | 0.73 | 0.66, 0.80 | 0.00 |  | 0.27 | 0.20, 0.34 | 1.03 | 0.87, 1.20 |
| ADE | 5.6 | 3 | 1.55 | 0.133 | 0.98 | 0.98 | 0.07 | 0.09 | 1989 | 0.64 | 0.09, 1.19 | 0.09 | -0.48, 0.66 | 0.27 | 0.20, 0.34 | 1.03 | 0.87, 1.19 |
| **AE** | **5.7** | **4** | **1.54** | **0.220** | **0.98** | **0.99** | **0.05** | **0.09** | **1983** | **0.73** | **0.66, 0.80** | **-** |  | **0.27** | **0.20, 0.34** | **1.03** | **0.87, 1.19** |
| CE | 33 | 4 | 0.87 | <0.001 | 0.72 | 0.86 | 0.20 | 0.12 | 2018 | - |  | 0.50 | 0.40, 0.60 | 0.50 | 0.41, 0.60 | 1.02 | 0.87, 1.17 |
| E | 104 | 5 | 1.41 | <0.001 | 0.06 | 0.62 | 0.33 | 0.26 | 2115 | - |  | - |  | 1.00 |  | 1.02 | 0.87, 1.17 |
| DunedinPoAm | | | | | | | | | | | | | | | | | |
| ACE | 1.3 | 3 | 1.12 | 0.722 | 1.00 | 1.02 | 0.00 | 0.04 | 2003 | 0.52 | 0.20, 0.85 | 0.09 | -0.20, 0.37 | 0.39 | 0.30, 0.48 | 0.98 | 0.86, 1.11 |
| ADE | 1.2 | 3 | 1.60 | 0.746 | 1.00 | 1.02 | 0.00 | 0.04 | 2003 | 0.62 | 0.53, 0.70 | 0.00 |  | 0.38 | 0.30, 0.47 | 0.98 | 0.86, 1.10 |
| **AE** | **1.6** | **4** | **1.20** | **0.802** | **1.00** | **1.02** | **0.00** | **0.04** | **1997** | **0.62** | **0.53, 0.70** | - |  | **0.38** | **0.30, 0.47** | **0.98** | **0.86, 1.10** |
| CE | 12.7 | 4 | 1.10 | 0.013 | 0.88 | 0.94 | 0.11 | 0.07 | 2009 | - |  | 0.45 | 0.36, 0.55 | 0.55 | 0.45, 0.64 | 0.98 | 0.86, 1.10 |
| E | 85.1 | 5 | 1.15 | <0.001 | 0.00 | 0.55 | 0.30 | 0.22 | 2087 | - |  | - |  | 1.00 |  | 0.98 | 0.86, 1.10 |
| DunedinPACE | | | | | | | | | | | | | | | | | |
| ACE | 4.1 | 3 | 2.19 | 0.256 | 0.99 | 0.99 | 0.04 | 0.01 | 2021 | 0.68 | 0.60, 0.76 | 0.00 |  | 0.32 | 0.24, 0.40 | 1.04 | 0.86, 1.23 |
| ADE | 3.6 | 3 | 1.79 | 0.301 | 0.99 | 1.00 | 0.03 | 0.08 | 2019 | 0.32 | -0.20, 0.85 | 0.36 | -0.18, 0.93 | 0.30 | 0.22, 0.38 | 1.03 | 0.86, 1.21 |
| **AE** | **5.4** | **4** | **1.64** | **0.248** | **0.98** | **0.99** | **0.04** | **0.09** | **2015** | **0.68** | **0.60, 0.76** | - |  | **0.32** | **0.24, 0.40** | **1.04** | **0.86, 1.23** |
| CE | 29 | 4 | 1.50 | <0.001 | 0.69 | 0.85 | 0.19 | 0.12 | 2050 | - |  | 0.42 | 0.31, 0.53 | 0.58 | 0.47, 0.69 | 1.02 | 0.86, 1.19 |
| E | 76.4 | 5 | 1.54 | <0.001 | 0.13 | 0.65 | 0.28 | 0.23 | 2118 | - |  | - |  | 1.00 |  | 1.02 | 0.86, 1.19 |
| Note. The epigenetic aging measures were adjusted for sex, age and baseline pubertal development prior to analysis; SC, scaling correction | | | | | | | | | | | | | | | | | |

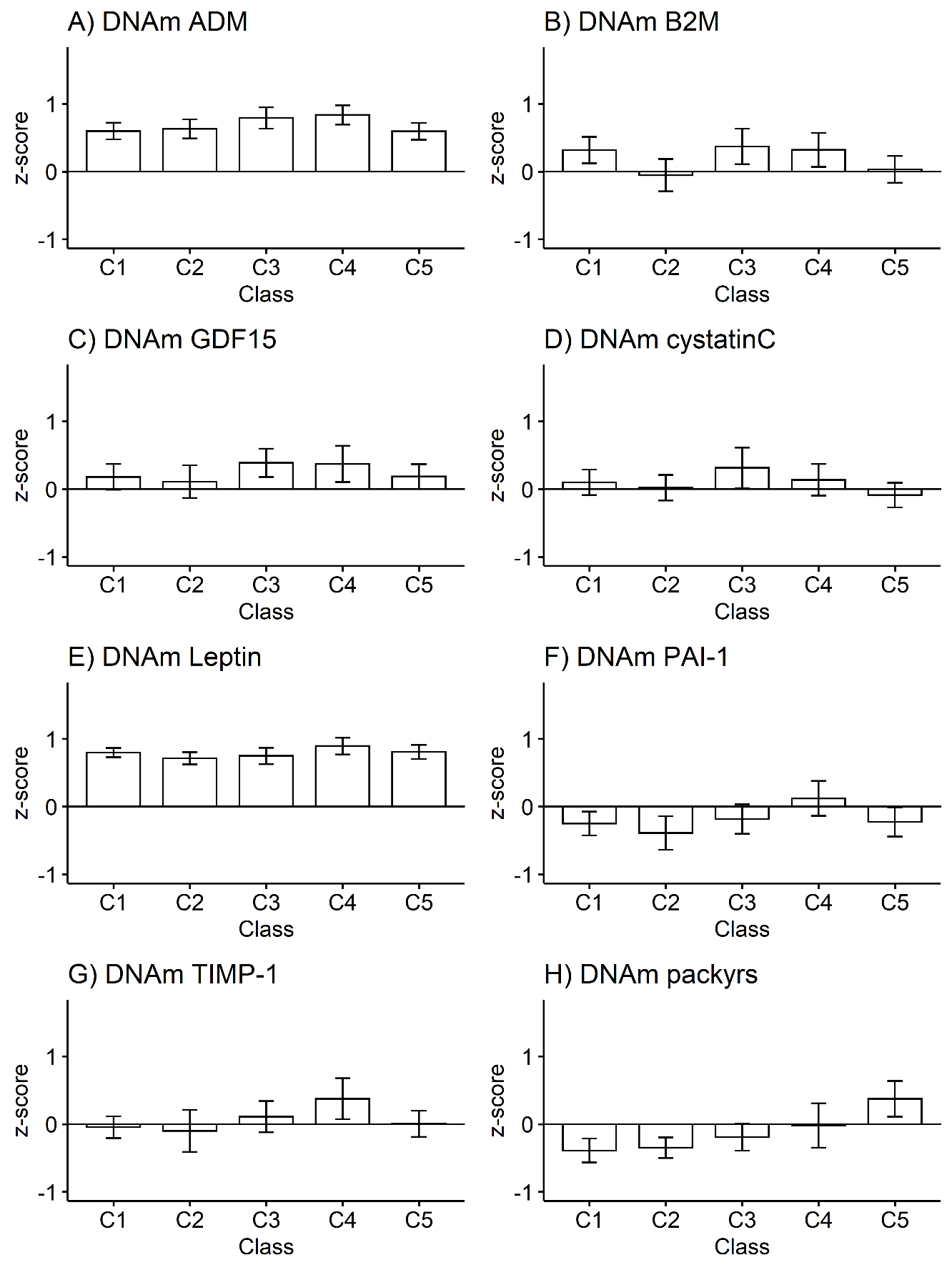

Figure S1. DNA methylation (DNAm)-based plasma proteins and smoking pack-years according to the adolescent lifestyle behavior patterns (n = 824): A) DNAm adrenomedullin (ADM), B) DNAm beta-2 microglobulin (B2M), C) DNAm growth differentiation factor (GDF15), D) DNAm cystatin C, E) DNAm leptin, F) DNAm plasminogen activation inhibitor 1 (PAI-1), DNAm tissue inhibitor metalloproteinase 1 (TIMP-1), and H) DNAm smoking pack-years (packyrs). Means and 95% confidence intervals are presented. C1 = the class with the healthiest lifestyle pattern, C2 = the class with low-normal BMI, C3 = the class with healthy lifestyle and high-normal BMI, C4 = the class with high BMI, C5 = the class with the unhealthiest lifestyle pattern. The model was controlled for sex (female), age and baseline pubertal development.

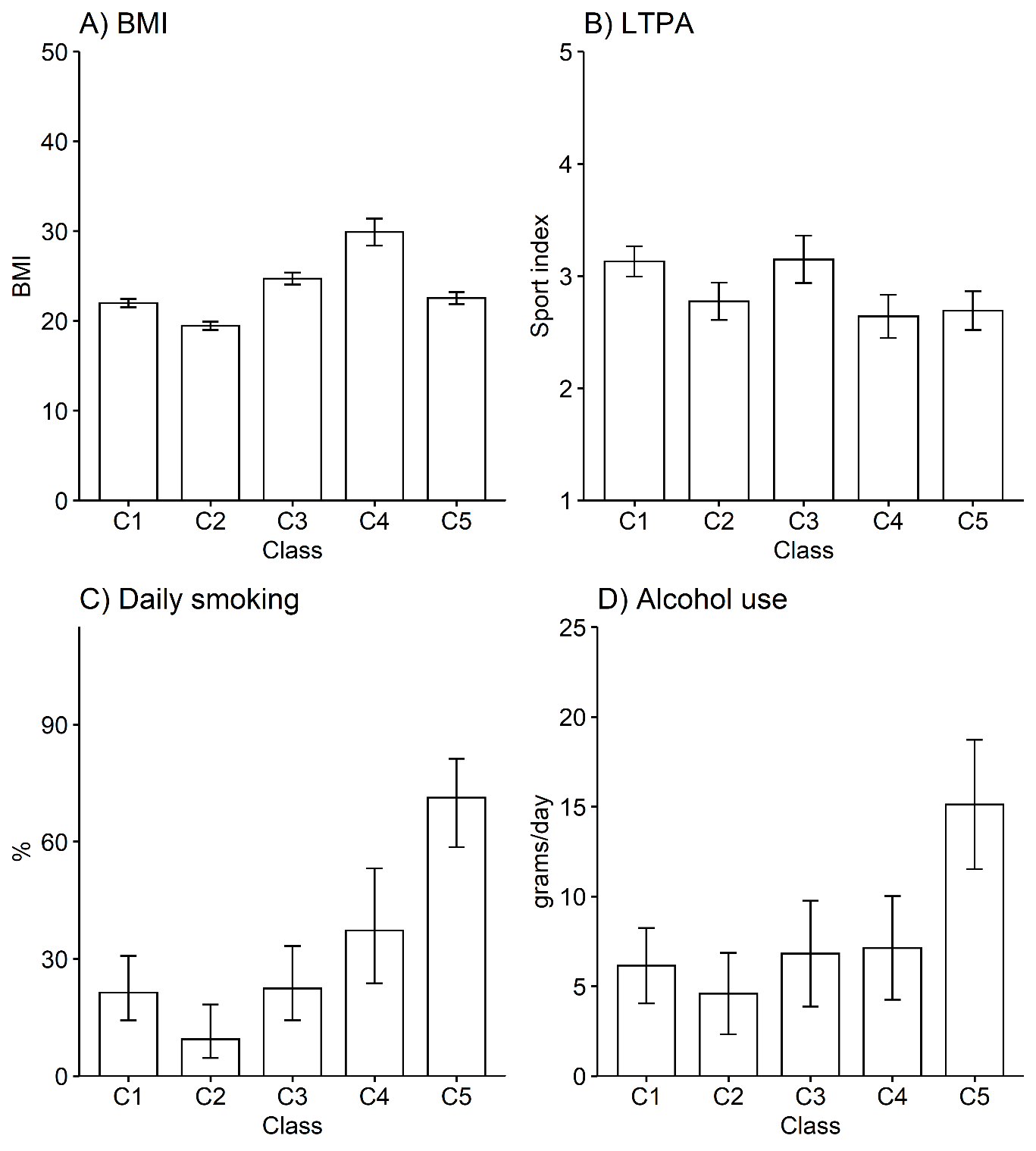

Figure S2. Lifestyle-related factors in adulthood according to the adolescent lifestyle behavior classes (n = 824): A) body mass index (BMI), B) leisure-time physical activity (LTPA), C) prevalence of daily smokers, and D) alcohol use. Means and 95% confidence intervals are presented. C1 = the class with the healthiest lifestyle, C2 = the class with low-normal BMI, C3 = the class with healthy lifestyle and high-normal BMI, C4 = the class with high BMI, C5 = the class with the unhealthiest lifestyle pattern. The model was controlled for sex (female), age and baseline pubertal development.
